# A two-stage super learner for healthcare expenditures

**DOI:** 10.1101/2021.03.26.21254428

**Authors:** Ziyue Wu, Seth Berkowitz, Patrick Heagerty, David Benkeser

## Abstract

**Objective:** To improve the estimation of healthcare expenditures by introducing a novel estimation method that is well-suited to situations where data exhibit strong skewness and zero-inflation.

**Data Sources:** Simulations, and two sources of real-world data: the 2016-2017 Medical Expenditure Panel Survey (MEPS) and the Back Pain Outcomes using Longitudinal Data (BOLD) datasets.

**Study Design:** Super learner is an ensemble machine learning approach that can combine several algorithms in order to improve estimation. We propose a two-stage super learner that is well suited for use with healthcare expenditure data by separately estimating the probability of any healthcare expenditure and the mean amount of healthcare expenditure conditional on having healthcare expenditures. These estimates can be combined to yield a single estimate of expenditures for each observation. The method can flexibly incorporate a range of individual estimation approaches for each stage of estimation, including both regression-based approaches and machine learning algorithms such as random forests. We compare the performance of the proposed two-stage super learner with a one-stage super learner, and with multiple individual algorithms for estimation of healthcare cost under a broad range of data settings in simulated and real data. The predictive performance of alternative strategies was compared using Mean Squared Error and R^2^.

**Principal Findings:** Our results indicate that the two-stage super learner has better performance compared with a one-stage super learner and individual algorithms, for healthcare cost estimation under a wide variety of settings in both simulations and empirical analyses. The improvement of the two-stage super learner over the one-stage super learner was particularly evident in settings when zero-inflation is high.

**Conclusions:** The two-stage super learner provides researchers an effective approach for healthcare cost analyses in environments where they cannot know the best single algorithm a priori.

## Introduction

The study of healthcare expenditures is a key component of health services research with common applications including estimating future expenditures, expenditures related to particular health conditions, and effects of interventions on expenditures [1]. However, statistical modeling of healthcare expenditures is often challenging for two reasons: individuals without health care utilization can lead to zero inflation, and very high expenditures in a small number of individuals can lead to large positive skewness [2–3]. In the United States, Berk & Monheit [4] report that in a typical year, 10% or more of individuals incur no healthcare expenditures, while 5% of the population accounts for the majority of health expenditures.

Many methods have been proposed to deal with these challenges [3]. A common approach involves regression of log-transformed cost using ordinary least squares. Duan’s smearing estimator [5] can be used to map estimates from the log-scale back to the original scale. Other approaches include non-Gaussian generalized linear models (GLM) [2, 5, 6], accelerated failure time (AFT) models [7], hazard-based models [8–10], and quantile-based models [11]. To explicitly account for zero-inflation, researchers often use two-part models [12]. In this approach, separate models are specified for the probability of any cost, and for the mean of costs amongst individuals with positive costs. Two-part GLMs with a logit link for the binary component and a Gamma distribution with log-link for the continuous component are common in practice [13–15].

All of these methods can work well when the outcome distribution meets the corresponding model assumptions. However, practitioners are often unaware of whether this is the case for a given data set and unfortunately, there is no “one-size-fits-all” approach [16]. Moreover, many common methods rely on parametric regression models, which are commonly mis-specified in practice, and can introduce bias in the results. This has led to increased interest in machine learning techniques, which use more flexible approaches to learn about relationships in data, thereby providing, in many cases, more accurate cost predictions [17]. Specifically, penalization and ensemble approaches have been applied for modeling healthcare expenditures [18–21].

In this work, we consider super learning, an ensemble method that combines a range of candidate models also known as model stacking, for application to healthcare expenditures [22–25]. Super learning entails using cross-validation to learn the optimal combination of a collection of candidate methods. This collection could include parametric and machine learning-based algorithms. Super learning has shown benefits over a single method in several healthcare studies [26–28] including prediction of expenditures in the context of plan payment risk adjustment [19].

However, no past research on super learning has focused specifically on zero-inflated data.

In this work, we propose a two-stage super learner. We define a set of candidate methods for predicting the presence of any costs, as well as for the positive portion of the cost distribution. The full super learner library consists of all pairwise combinations of the methods for each stage. We provide an efficient implement of the two-stage super learner with tailoring for challenges associated with healthcare cost data. We compare the two-stage super learner with the regular, one-stage super learner, along with individual methods commonly used in studies of healthcare expenditures using Monte Carlo simulations under various data generating processes. In addition, we analyze data from the 2016-2017 Medical Expenditure Panel Survey (MEPS) [29, 30] and Back pain Outcomes using Longitudinal Data (BOLD) project [31]. In both cases, the two-stage super learner improved performance over existing approaches.

## Methods

### Two-stage super learner

#### Two-part model

The two-part model consists of a binary model for the probability of outcome being positive and a regression model applied to the positive subsample. Let Y denote the healthcare cost, and X denote a potentially high-dimensional vector of predictors.

Motivation for a two-part model is that the mean, *E*[*Y*|*X*], can be written as:

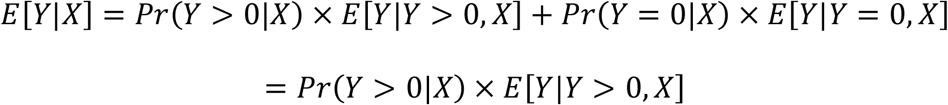

The two pieces of this equation can be estimated separately. The first piece, *Pr*(*Y* > 0 | *X*) is commonly modeled using logistic regression, although in principle any regression suitable for a binary outcome could be used. The second component *E*[*Y*|*Y* > 0, *X*] is commonly modeled using Gamma-family GLMs with log link functions to account for high skewness, though again any suitable regression could be used. Given the dizzying array of possible regression approaches for each component of this model, it could be quite useful for practitioners to have both a formal framework for selecting amongst the many choices and a general recommendation on an optimal strategy when the goal is to obtain high prediction accuracy.

#### Super Learning

Super learning provides a formal means of selecting a combination of algorithms that best fits the true regression function. Here, the notion of “best” refers to a cross-validated risk criterion. Suppose we have a dataset of independent observations on n individuals, *O*_*i*_ = (*Y*_*i*_, *X*_*i*_), *i* = 1, …, *n*, where *Y* is the outcome of interest and *X* is a set of covariates. Suppose we have access to a candidate prediction function 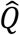, e.g.,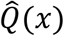 that provides a prediction for a new observation *x* based on a fitted two-stage model. We introduce the notion of the *risk of* 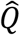, which provides a global summary of how well 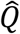 predicts outcomes *Y* based on covariates *X*. Often [32] we rely on risk measures that can be expressed as the *average discrepancy* between *Y*and the prediction made by 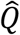. In other words, given a regression 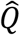, we can define a *loss function* 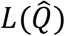, that takes as input a particular data point (*x, y*) and returns a real number measuring the discrepancy between 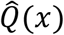 and *y*. A larger value of the loss indicates a further gap between prediction and truth. Common loss functions for cost data include mean squared error (MSE) for mean cost estimation, mean absolute error (MAE) for median cost estimation and “check function” [33] for quantile cost estimation (e.g. 90^th^ percentile of cost). In this study, we explicitly consider the squared-error loss function 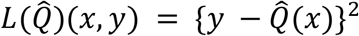 and consider mean squared error (MSE), 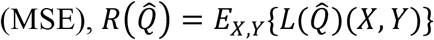, as our risk criterion. Given a risk criterion, we can define the optimal prediction function, say *Q*_0_ as the function that minimizes risk over all possible prediction functions.

The performance optimization problem can be equivalently defined as a statistical estimation problem. For example, it is straightforward to show that for squared error loss, *Q*_0_(*X*) = *E*(*Y*|*X*). Thus, the task of learning the optimal (with respect to MSE) prediction function is equivalent to the task of estimating the conditional mean of cost given covariates. Super learner provides one objective and flexible strategy for accomplishing this task.

In a given problem, there are often many different individual approaches to developing a prediction function. In super learning, we call each approach an *algorithm* and refer to a pre-specified collection of algorithms as a *library*. Here, “algorithm” is used in a general sense as any means of mapping a given data set into a prediction function. We use *training* to refer to the process of applying an algorithm to data. Examples of algorithms include (i) fitting ordinary least squares regression and returning the linear predictor; (ii) performing variable screening based on a univariate significance threshold, *then* applying ordinary least squares regression; (iii) training a random forest, where tuning parameters are selected via cross-validation. The super learner library should be, to the greatest extent possible, informed by subject-area expertise, but can utilize data-driven, machine learning approaches as well [34].

Suppose we have *K* potential algorithms. The implementation of super learner involves using cross-validation to determine an ensemble of these algorithms. The process can be implemented as follows.

1. Train each algorithm in the library on the entire dataset yielding prediction functions 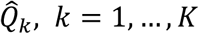.
2. Randomly split the data into *V*mutually exclusive and exhaustive blocks of approximately equal size. For *v* = 1, …, *V*, define the *v*-th block as the *v*-th validation sample, and the remaining *V* − 1 blocks the *v*-th training sample.
3. For *v* = 1, …, *V*, train each of the *K* algorithms using the *v*-th training sample yielding prediction functions 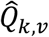. For each observation *X*_*i*_ in the *v*-th validation sample obtain the prediction 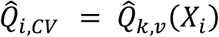.
4. Propose an ensemble of algorithms in the library indexed by *K*-length-weight vector *α*, e.g.,

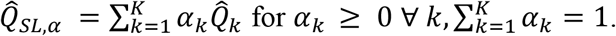
5. Find the weights 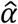 that minimize the cross-validated mean squared error of the ensemble over all possible values of *α*, e.g.,

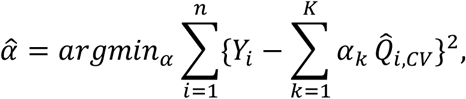

where the minimum is taken over all 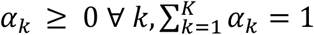.
6. The super learner is 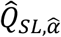.

In step 4, many choices of ensembles could be considered. One particular choice is to build the ensemble from a set of weight vectors that each assigns a weight of 1 to a particular algorithm and 0 weight to all others. The choice of assigning weight of 1 to a particular algorithm is known as the cross-validation selector or discrete super learner since it represents the individual algorithm with the lowest cross-validated risk.

Theoretical guarantees [25, 35, 36] state that in large samples the super learner should predict costs essentially as well as or better than the best-performing single algorithm amongst the *K* considered. Because the super learner is only capable of predicting as well as the best amongst the *K* algorithms used in its construction, this theory suggests that it is advantageous to consider a diverse mixture of algorithms when constructing a super learner. In the next section, we propose a strategy for obtaining a large and varied collection of modeling approaches in the context of zero-inflated data.

#### Two-stage Super Learner

We propose a two-stage Super Learner model, wherein we propose a library for *Pr*(*Y* > 0 | *X*) *and for E*[*Y* | *Y* > 0, *X*]. The overall two-stage super learner library consists of all pairwise combinations of these two models. Assuming the stage-1 library includes *K*_1_ algorithms and the stage-2 library includes *K*_2_ algorithms, then the two-stage super learner’s “whole library” would contain *K*_1_ × *K*_2_ candidate algorithms with each one representing a specific combination of algorithms from the stage-1 and stage-2.

The implementation of the two-stage super learner is similar to that of the super learner described above. The key difference is that each of our proposed candidate algorithms consists of two constituent algorithms: one for *Pr*(*Y* > 0 | *X*) and one for *E*[*Y* | *Y* > 0, *X*]. A prediction of *Y* is obtained as the product of the predictions made by each trained algorithm. Beyond this modification in how predictions are obtained, our approach is implemented as described above and the theoretical results governing the behavior of the original super learner immediately apply to our approach.

Our approach is implemented in a freely available R package. In addition to the two-stage learning approach, our software handles additional challenges associated with healthcare expenditure data. In particular, we implement a novel cross-validation approach that ensures an approximately equal split of zeros and outliers over the folds. The package also provides a modified quadratic programming approach to calculate super learner weights in the presence of very large residual values (Appendix-A).

#### One-stage Super Learner

The standard one-stage super learner is similar to the two-stage super learner, except that it does not split the estimation problem into two parts. When zero inflation is high, this may lead to decreased performance. However, if zero inflation is not prominent in a particular dataset, it may not be necessary to estimate the probability of zero expenditures, and thus one-stage super learner may perform well. The one-stage super learner can be computationally faster than the two-stage super learner.

Since the one-stage super learner has not been widely used for healthcare cost estimation, an additional objective of this study was to examine the performance of one-stage super learner in various situations a healthcare cost analyst may encounter.

#### Monte Carlo Simulations

We evaluated the relative performance of the two-stage super learner compared to common approaches across a wide range of settings using Monte Carlo simulations. In particular, we studied how the two-stage super learner behaved in comparison with one-stage super learner and individual algorithms when varying the sample size, zero-inflation amount, non-zero distribution and complexity in the cost data.

#### Data generating process

For simulation study, categorical and continuous predictors were generated to include as many scenarios as possible and approximate covariates seen in real data.

Specifically, predictors were simulated as follows:

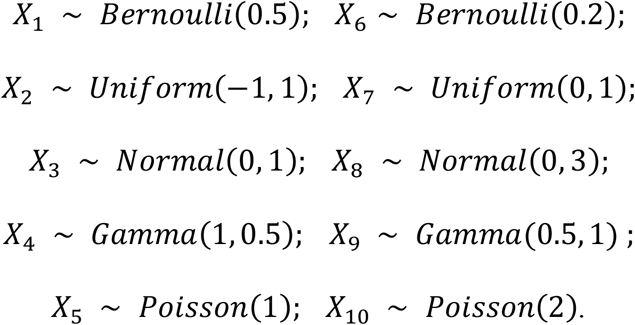

Variables *X*_1_, …, *X*_5_ impacted the distribution of costs, while the others were noise. To assess how the sample size, zero percentage, non-zero distribution, and data complexity affects the estimations, we varied four aspects of the data generating process: sample size (small (500) vs. large (2000)), zero inflation percentage (low (5%) vs. high (70%)), distribution of non-zero costs (Log-normal, Gamma, Tweedie, Mixture), and data complexity (two-way interactions among covariates, Yes and No). The combination of the four settings above results in 32 scenarios. For each scenario, we analyzed 1000 simulated data sets.

Costs were simulated using a two-stage procedure to allow for point mass at zero. In the first stage, we drew a random variable *Z* from a Bernoulli distribution with the probability of zero determined by a logistic model (Appendix-B). If *Z* = 0, then we set costs to zero. If *Z* = 1, we drew the observed costs from one of the four different non-zero distributions listed above (Appendix-C & Appendix-Figure 1).

**Figure 1:**
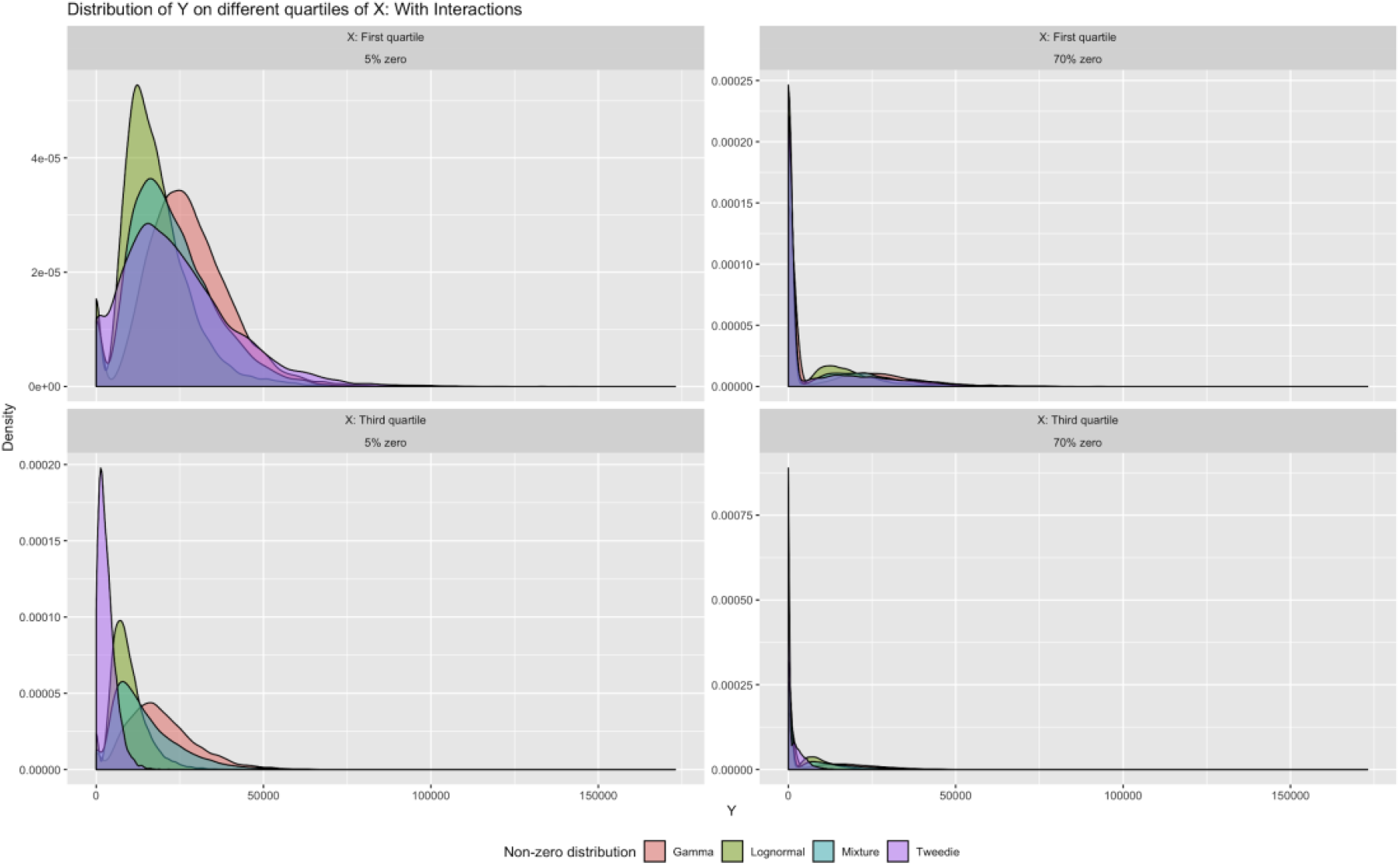
Distributions of Y given X set to first & third quartile values. Figure 1 illustrates the diversity of the distributions studied in our simulation. In the top row, we show distributions for the interaction scenario under low (left column) and high (right column) zero inflation when all predictors are set equal to their respective first quartile values. The bottom row shows the same, but with predictors set to their respective third quartile values.

#### Prediction algorithms

A previous study [37] indicated that the choice of the stage-2 model may alter results more dramatically than stage-1. Accordingly, we focused our two-stage super learner towards diversity in algorithms for stage-2 by including 10 algorithms in the stage-2 model, while only including 3 algorithms in the stage-1 model. Additionally, we were interested in how the performance of a two-stage super learner compared to that of a standard super learner, thus we also fit a standard super learner with a library of 8 single-stage algorithms. Hence, we refer to this algorithm as the *one-stage* super learner. The algorithms considered in the simulation study include a mixture of machine learning algorithms such as random forests and parametric regressions such as GLM with Gamma distribution and log link (Appendix-Table 3). We compared the performance of each of 38 algorithms (30 combinations of two-stage algorithms + 8 individual algorithms), the one-stage super learner, the two-stage super learner built from 38 algorithms, and the *discrete super learner*, or the single algorithm in the two-stage super learner library with lowest cross-validated risk as comparisons to the two-stage super learner. For each two-stage super learner, we used *V* = 10 folds for cross-validation.

**Table 3.** MSE, relative MSE, R^2^, and Relative Efficiency (RE) for top 10 algorithms in modeling four spine-related RVUs

| Algorithm | MSE | Relative<br>MSE | $R^2$ | RE |
| --- | --- | --- | --- | --- |
| Spine-related RVUs (5% zero) |  |  |  |  |
| Two-stage Super Learner | 6275.351 | 1.0000 | 0.0621 | 1.0000 |
| One-stage Super Learner | 6307.790 | 1.0052 | 0.0611 | 0.9839 |
| Discrete Super Learner | 6316.867 | 1.0066 | 0.0608 | 0.9791 |
| Single: RF | 6320.680 | 1.0072 | 0.0607 | 0.9775 |
| S1: RF + S2: RF | 6336.996 | 1.0098 | 0.0600 | 0.9662 |
| S1: GLM + S2: RF | 6344.708 | 1.0111 | 0.0597 | 0.9614 |
| S1: Lasso + S2: RF | 6344.743 | 1.0111 | 0.0597 | 0.9614 |
| S1: RF + S2: Lasso | 6394.274 | 1.0190 | 0.0579 | 0.9324 |
| S1: Lasso + S2: Lasso | 6395.829 | 1.0192 | 0.0578 | 0.9308 |
| S1: GLM + S2: Lasso | 6396.604 | 1.0193 | 0.0578 | 0.9308 |
| Spine-related imaging RVUs (55% zero) |  |  |  |  |
| Two-stage Super Learner | 55.4923 | 1.0000 | 0.0879 | 1.0000 |
| Discrete Super Learner | 55.6790 | 1.0034 | 0.0861 | 0.9784 |
| S1: RF + S2: GBM | 55.7779 | 1.0051 | 0.0849 | 0.9659 |
| S1: RF + S2: Lasso | 55.7976 | 1.0055 | 0.0845 | 0.9613 |
| S1: RF + S2: RF | 55.8768 | 1.0069 | 0.0832 | 0.9465 |
| One-stage Super Learner | 55.8939 | 1.0072 | 0.0830 | 0.9443 |
| S1: RF + S2: GLM-Gamma-Identity | 55.9810 | 1.0088 | 0.0815 | 0.9272 |
| S1: RF + S2: GLM-Gamma-Log | 55.9972 | 1.0091 | 0.0813 | 0.9249 |
| S1: RF + S2: Log-OLS smearing | 56.0175 | 1.0095 | 0.0809 | 0.9204 |
| S1: GLM + S2: Lasso | 56.0735 | 1.0105 | 0.0800 | 0.9101 |
| Spine-related physical therapy RVUs (85% zero) |  |  |  |  |
| Two-stage Super Learner | 2.4964 | 1.0000 | 0.2778 | 1.0000 |
| Discrete Super Learner | 2.5141 | 1.0071 | 0.2671 | 0.9615 |
| S1: Lasso + S2: Log-OLS smearing | 2.5213 | 1.0100 | 0.2624 | 0.9446 |
| S1: GLM + S2: Log-OLS smearing | 2.5233 | 1.0108 | 0.2618 | 0.9424 |
| S1: Lasso + S2: GLM-Gamma-Identity | 2.5260 | 1.0119 | 0.2610 | 0.9395 |
| S1: GLM + S2: GLM-Gamma-Identity | 2.5265 | 1.0121 | 0.2609 | 0.9392 |
| S1: GLM + S2: Lasso | 2.5270 | 1.0123 | 0.2607 | 0.9384 |
| S1: Lasso + S2: Lasso | 2.5294 | 1.0132 | 0.2600 | 0.9359 |
| S1: Lasso + S2: GBM | 2.5298 | 1.0134 | 0.2599 | 0.9356 |
| S1: GLM + S2: GBM | 2.5314 | 1.0140 | 0.2594 | 0.9338 |
| One-stage Super Learner | 2.5332 | 1.0147 | 0.2587 | 0.9312 |
| Spine-related injection RVUs (91% zero) |  |  |  |  |
| Two-stage Super Learner | 17.1029 | 1.0000 | 0.1235 | 1.0000 |
| Discrete Super Learner | 17.2228 | 1.0070 | 0.1183 | 0.9579 |
| S1: Lasso + S2: Lasso | 17.2480 | 1.0085 | 0.1172 | 0.9490 |
| S1: Lasso + S2: GLM-Gamma-Identity | 17.2543 | 1.0089 | 0.1169 | 0.9466 |
| S1: GLM + S2: Lasso | 17.2564 | 1.0090 | 0.1168 | 0.9457 |
| S1: GLM + S2: Log-OLS smearing | 17.2571 | 1.0090 | 0.1167 | 0.9449 |
| S1: GLM + S2: GLM-Gamma-Identity | 17.2575 | 1.0091 | 0.1167 | 0.9449 |
| S1: Lasso + S2: GLM-Gamma-Log | 17.2596 | 1.0092 | 0.1165 | 0.9433 |
| S1: Lasso + S2: Log-OLS smearing | 17.2605 | 1.0092 | 0.1164 | 0.9425 |
| S1: GLM + S2: GLM-Gamma-Log | 17.2637 | 1.0094 | 0.1161 | 0.9401 |
| One-stage Super Learner | 17.2993 | 1.0115 | 0.1138 | 0.9215 |

MSE, relative MSE, $R^2$ , and RE are calculated using 10-fold cross-validation. S1 refers to stage-1 and S2 refers to stage-2. RF refers to Random forest. GLM in S1 refers to logistic regression and Lasso in S1 refers to logistic Lasso regression. GLM-Gamma-Log refers to GLM with Gamma distribution and Log link function. Log-OLS smearing refers to logarithmic OLS with smearing retransformation. GBM refers to gradient boosting machine. Lower MSE and relative MSE indicate better performance. Greater $R^2$ and RE indicate better performance. For spine-related physical therapy RVUs and spine-related injection RVUs, the one-stage super learner is not among the top 10 algorithms, we still list results for comparison.

#### Evaluation metrics

Performances of prediction algorithms (individual algorithms, one-stage super learner, discrete super learner, and two-stage super learner) were evaluated on an independent test set of size *n*_*test*_. Let *y*_*i*_ be the true outcome values on the test set, ŷ_*i*_ be the predicted outcome values from certain algorithms on the test set and 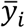 be the mean of *y*_*i*_, we use the following metrics for evaluation:

Mean Squared Error (MSE):

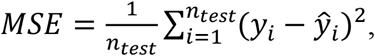

Relative MSE:

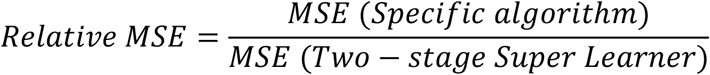

Coefficient of determination (R^2^):

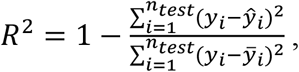

Relative Efficiency (RE):

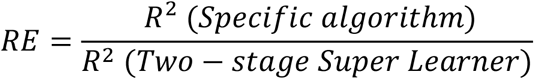

### Real-world data

#### MEPS Data

We analyzed the Medical Expenditure Panel Survey (MEPS) data from 2016 to 2017 [29, 30]. MEPS is a national survey on the financing and use of medical care of families and individuals, their medical providers, and employers across the United States. Participating households were drawn from a nationally representative subsample and provided demographic information, health status, self-reported medical conditions, medical expenditure and utilization, health insurance coverage, and access to care for medical events. For some individuals, self-reported medical expenditures are supplemented with information from medical providers and insurers.

We used the longitudinal data of MEPS 2016-2017, with 2016 MEPS used as a training sample and the same individuals in the 2017 MEPS as a testing sample. We developed a prediction model for total annual healthcare expenditures based on 14 covariates that included demographics, medical conditions, and insurance characteristics (Appendix-Table 7). Total annual healthcare expenditures include out-of-pocket payments and third-party payments from all sources but exclude insurance premiums. The sample included only adults and excluded observations with missing data. The final sample contained 10,925 observations for training and 10,815 observations for testing. We used the same library of algorithms as the simulations and ten-fold cross-validation to build the two-stage super learner (Appendix-Table 3).

#### BOLD Data

The Back Pain Outcomes using Longitudinal Data (BOLD) project is a large, community-based registry of patients aged 65 years and older who presented with primary care visits for a new episode of back pain from March 2011 to March 2013 [31]. The BOLD data are derived from self-reported questionnaires and electronic health records (EHR). Expenditures were calculated as total relative value units (RVUs), a measure of value used in the US Medicare reimbursement formula for physician services [38].

In this study, we considered 4 spine-related total annual RVUs as outcomes, including 1) total spine-related RVUs; 2) spine-related physical therapy RVUs; 3) spine-related injection RVUs; 4) spine-related imaging RVUs. We developed separate prediction models of each outcome based on 24 covariates from baseline patient questionnaires and EHR, see Appendix-D & Appendix-Table 9 for details. The final sample contained 4,397 observations. Some algorithms used in our simulation study failed to converge on the BOLD data, necessitating the use of a slightly different library of algorithms for this part of the study (Appendix-Table 10). To evaluate the models, the data were randomly partitioned into ten distinct folds. Every single algorithm, one-stage super learner, discrete super learner and the two-stage super learner were trained using nine of the folds and predictions were obtained using the remaining fold. The performance was estimated as the MSE of the predictions on the validation fold, averaged over the ten validation folds.

## Results

### Monte Carlo Simulation

On average, across all settings, the two-stage super learner had the smallest MSE (Table 1), alongside discrete super learner and one-stage super learner. See Appendix-Table 4 & 5 for results under other evaluation metrics. Overall, the discrete super learner (the approach that used cross-validation to select the single best algorithm from the two-stage super learner library for each situation, as opposed to using a weighted average of all algorithms) was the second-best-performing algorithm, while the one-stage super learner was the third. The overall MSE improvements of two-stage super learner over discrete learner, one-stage super learner and the best single algorithm were about 1.9%, 4.0% and 9.1%, respectively. Other algorithms with favorable performances included combinations of the Lasso model at stage-1 with GLM (log-link & gamma distribution) at stage-2 and Zero-inflated Negative-Binomial (ZINB) model. The overall Interquartile Range (IQR) of MSE for the two-stage super learner is smaller compared to other algorithms, indicating a robust prediction performance under different data settings. The improvement of the two-stage super learner over the one-stage super learner was more evident under high zero-inflation (70% zero) with a MSE improvement of 9.7%. This was in line with expectations as using a two-stage model is likely to be more beneficial the greater the point mass at zero. When zero inflation was small (5% zero), the one-stage super learner was superior and had the smallest MSE, but the two-stage super learner was the second best-performing algorithm and outperformed the best single algorithm (ZINB) with a MSE improvement of 4.1%.

**Table 1.** Average and IQR of MSE for algorithms across 32 data generating processes

| Algorithm | MSE (mean + IQR) ($10^8$) | | |
| --- | --- | --- | --- |
|  | Overall | Low zero-inflation | High zero-inflation |
| Two-stage Super Learner | 3.239 [1.354, 4.203] | 3.437 [1.329, 4.475] | 3.041 [1.394, 4.043] |
| Discrete Super Learner | 3.301 [1.364, 4.362] | 3.459 [1.272, 4.494] | 3.143 [1.483, 4.204] |
| One-stage Super Learner | 3.374 [1.482, 4.424] | 3.381 [1.398, 4.472] | 3.367 [1.692, 4.367] |
| S1: Lasso + S2: GLM-Gamma-Log<sup>†</sup> | 3.563 [1.511, 4.610] | 3.780 [1.446, 5.066] | 3.345 [1.549, 4.230] |
| S1: Lasso + S2: Quantile regression | 3.567 [1.502, 4.579] | 3.784 [1.447, 5.066] | 3.350 [1.546, 4.219] |
| S1: Lasso + S2: Log OLS-smearing | 3.570 [1.504, 4.614] | 3.786 [1.448, 5.066] | 3.354 [1.546, 4.232] |
| Zero-inflated Negative Binomial (ZINB)<sup>‡</sup> | 3.582 [1.526, 4.641] | 3.585 [1.363, 4.820] | 3.579 [1.641, 4.529] |
| S1: GLM + S2: GLM-Gamma-Log | 3.583 [1.511, 4.638] | 3.809 [1.451, 5.124] | 3.356 [1.543, 4.246] |
| S1: GLM + S2: Quantile regression | 3.589 [1.511, 4.606] | 3.818 [1.448, 5.140] | 3.359 [1.536, 4.227] |
| S1: GLM + S2: Log OLS-smearing | 3.590 [1.512, 4.640] | 3.819 [1.448, 5.134] | 3.361 [1.538, 4.234] |
| Zero-inflated Poisson (ZIP) | 3.601 [1.631, 4.751] | 3.613 [1.485, 4.919] | 3.589 [1.770, 4.655] |
| S1: Lasso + S2: Adaptive GLM | 3.784 [1.531, 4.804] | 3.905 [1.398, 5.017] | 3.663 [1.638, 4.623] |
| S1: GLM + S2: Adaptive GLM | 3.803 [1.524, 4.831] | 3.937 [1.406, 5.089] | 3.668 [1.632, 4.615] |
| S1: RF + S2: GLM-Gamma-Log | 4.381 [2.176, 5.453] | 4.604 [2.227, 5.890] | 4.157 [2.115, 5.002] |
| S1: RF + S2: Quantile regression | 4.385 [2.180, 5.396] | 4.609 [2.237, 5.855] | 4.160 [2.122, 4.973] |
| S1: RF + S2: Log OLS-smearing | 4.390 [2.185, 5.447] | 4.617 [2.232, 5.911] | 4.163 [2.130, 4.997] |
| S1: RF + S2: Adaptive GLM | 4.626 [2.204, 5.658] | 4.937 [2.261, 6.133] | 4.314 [2.176, 5.111] |
| S1: GLM + S2: RF | 4.898 [1.833, 7.055] | 5.550 [1.861, 8.202] | 4.246 [1.809, 6.232] |
| S1: Lasso + S2: RF | 4.900 [1.830, 7.055] | 5.556 [1.866, 8.217] | 4.244 [1.814, 6.224] |
| RF | 5.304 [2.270, 7.396] | 5.439 [2.033, 7.782] | 5.168 [2.592, 7.139] |
| S1: RF + S2: RF | 5.506 [2.262, 7.701] | 6.250 [2.256, 9.016] | 4.761 [2.264, 6.723] |
| S1: Lasso + S2: AFT (generalized Gamma) | 6.321 [2.014, 9.634] | 6.458 [1.949, 9.906] | 6.183 [2.044, 9.357] |
| S1: GLM + S2: AFT (generalized Gamma) | 6.339 [2.013, 9.650] | 6.488 [1.938, 9.930] | 6.190 [2.046, 9.359] |
| S1: RF + S2: AFT (generalized Gamma) | 7.049 [2.714, 10.650] | 7.332 [2.801, 11.014] | 6.765 [2.651, 10.081] |
| Tobit | 9.165 [3.129, 14.079] | 9.184 [2.920, 13.677] | 9.145 [3.283, 14.415] |
| S1: Lasso + S2: Lasso (OLS) | 9.295 [2.082, 15.509] | 9.742 [2.019, 15.715] | 8.848 [2.118, 15.241] |
| S1: GLM + S2: Lasso (OLS) | 9.313 [2.099, 15.523] | 9.776 [2.028, 15.844] | 8.850 [2.120, 15.275] |
| Lasso (OLS) | 9.677 [2.553, 15.509] | 9.710 [2.316, 15.273] | 9.643 [2.739, 15.754] |
| OLS | 9.703 [2.571, 15.539] | 9.737 [2.356, 15.304] | 9.668 [2.744, 15.771] |
| S1: RF + S2: Lasso (OLS) | 9.907 [2.366, 16.238] | 10.373 [2.292, 16.527] | 9.440 [2.395, 15.869] |
| S1: Lasso + S2: Adaptive hazard | 9.920 [2.463, 14.535] | 12.270 [2.782, 18.840] | 7.570 [2.275, 11.282] |
| S1: GLM + S2: Adaptive hazard | 9.933 [2.463, 14.568] | 12.292 [2.768, 18.995] | 7.574 [2.276, 11.294] |
| S1: RF + S2: Adaptive hazard | 10.226 [2.600, 14.994] | 12.504 [2.758, 19.367] | 7.947 [2.449, 11.819] |
| Tweedie | 11.273 [2.558, 16.989] | 12.155 [2.331, 16.723] | 10.391 [2.879, 17.323] |
| S1: GLM + S2: Cox hazard | 12.341 [2.134, 19.414] | 12.510 [1.958, 19.552] | 12.172 [2.302, 19.289] |
| S1: Lasso + S2: Cox hazard | 12.353 [2.142, 19.442] | 12.525 [1.973, 19.664] | 12.181 [2.303, 19.335] |
| S1: RF + S2: Cox hazard | 12.820 [2.461, 20.298] | 13.030 [2.314, 20.360] | 12.609 [2.600, 20.242] |
| S1: GLM + S2: GLM-Gamma-Identity | 13.249 [2.230, 22.589] | 13.351 [2.046, 22.284] | 13.147 [2.437, 22.696] |
| S1: Lasso + S2: GLM-Gamma-Identity | 13.263 [2.248, 22.583] | 13.369 [2.059, 22.267] | 13.157 [2.440, 22.713] |
| S1: RF + S2: GLM-Gamma-Identity | 13.717 [2.519, 22.934] | 13.867 [2.351, 22.812] | 13.567 [2.677, 23.096] |
| OLS intercept only (mean) | 17.512 [2.900, 28.989] | 17.560 [2.649, 28.778] | 17.464 [3.096, 29.118] |

<sup>†</sup>Top single algorithm under high zero-inflation. <sup>‡</sup>Top single algorithm under low zero-inflation. Lower MSE indicates better performance. Algorithms are presented in ascending order according to the overall mean MSE. S1 refers to stage-1 and S2 refers to stage-2. GLM in S1 refers to logistic regression and Lasso in S1 refers to logistic Lasso regression. RF refers to Random Forest. GLM-Gamma-Log refers to GLM with Gamma distribution and Log link function.

Appendix-Figure 2 presents more extensive results. Overall, we found that no single algorithm was best in all cases, and there are instances where a single algorithm provides the best performance. Practically, however, the analyst will not know which algorithm fits the data best a priori, which supports the idea of using a super learner approach. Across a large number of cases, the two-stage super learner was more robust than the individual algorithms and close to the best algorithm under all different data settings, while the performances of individual algorithms changed dramatically across different data settings.

**Figure 2:**
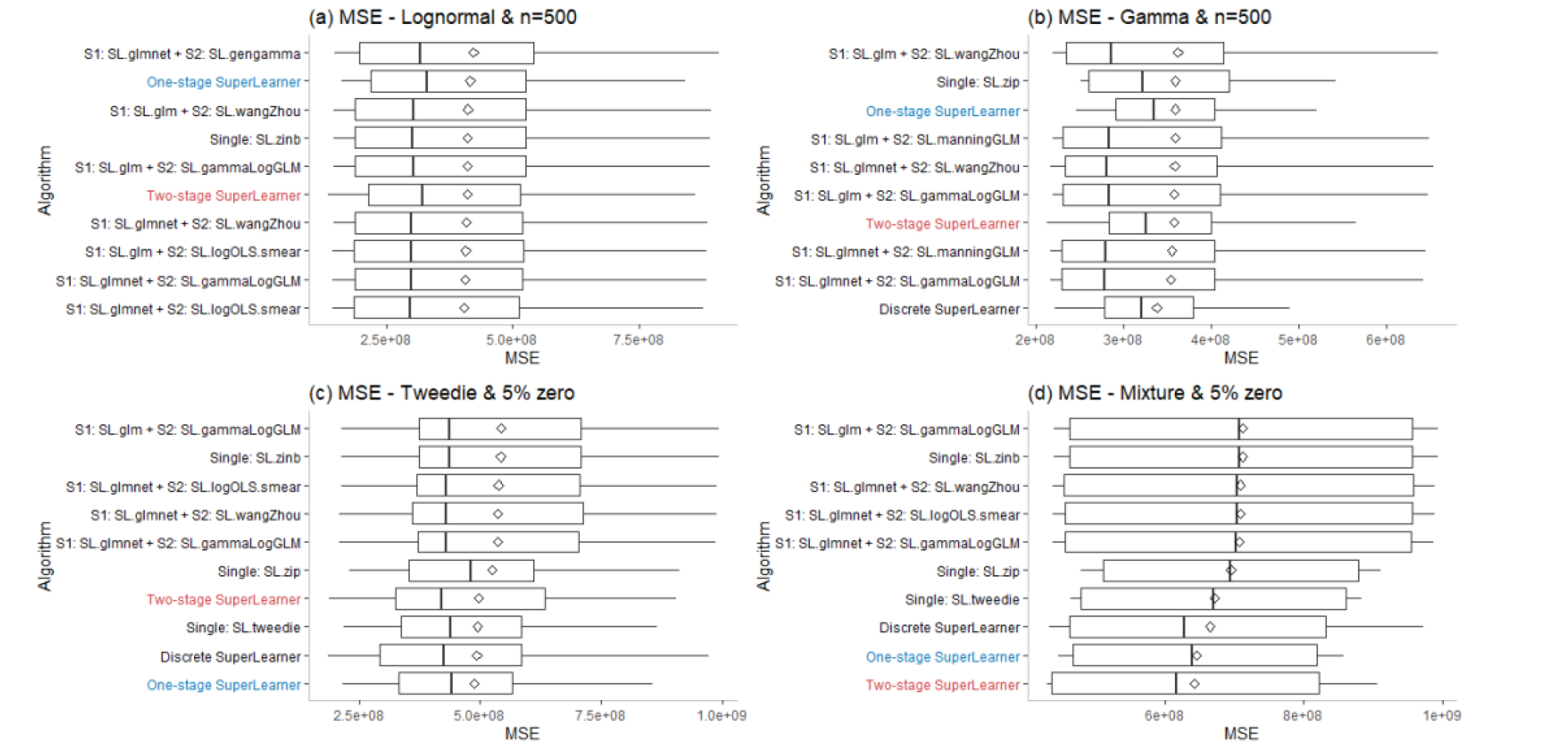
Boxplot of MSE for top 10 algorithms under 4 different data settings in Monte Carlo simulations. There are cases when the two-stage super learner was not best behaved as no single best model exists among all situations. Not surprisingly, log-OLS with smearing retransformation (figure 1a), gamma GLM with log link (figure 1b) and Tweedie model (figure 1c) had fairly good prediction performances and beaten the two-stage super learner under their corresponding data generating distributions since these estimators were able to well approximate the true underlying data structure. However, by aggregating diverse component model predictions into a final ‘meta prediction, the two-stage super learner was more robust than any component models and close to the best model under different data settings, while the performances of component models changed dramatically across different settings.

### Empirical analysis

#### MEPS

Distributions of total expenditures in 2016 & 2017 were both zero-inflated, with almost 20% of observations having zero expenditures, as well as highly skewed, with 2% of observations expenditures over $50,000 (Appendix-Table 6, 7 & Appendix-Figure 3).

**Figure 3:**
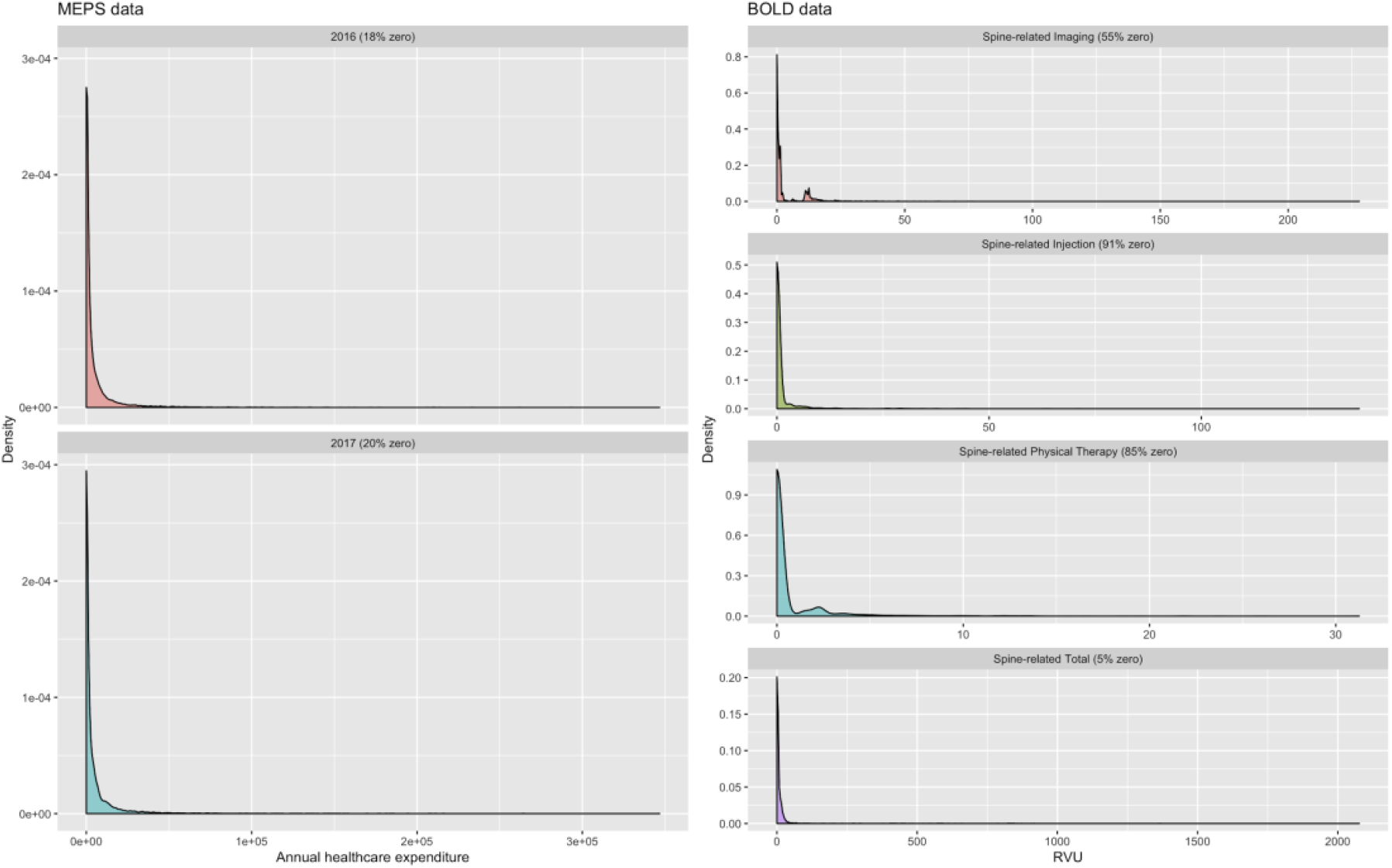
Distributions of outcomes in MEPS analysis & BOLD analysis Figure 3 illustrates the distributions of outcomes in MEPS analysis (left column) and BOLD analysis (right column). All the outcomes in the two analyses are highly skewed with a mass at zero and heavy upper tails.

The two-stage super learner performed better than all the individual algorithms and the one-stage super learner with the lowest test-set MSE (table 2). Specifically, for MSE, the improvements of the two-stage super learner over the best single algorithm and one-stage super learner were about 0.7% and 2.2%; while for R^2^, such improvements increased to 4.2% and 14.6%. The one-stage super learner was among the top 10 algorithms with MSE lower than any single-stage individual algorithm (table 2). Relative MSE for individual algorithms compared to the two-stage super learner ranged from 1.007 (Random Forest at both stages) to 1.192 (model with intercept only). Adding stage-1 for zero-inflation appears to improve the prediction as Random Forest and Lasso used at stage-2 of two-stage models were better than them being used solely in one-stage models. Choices of stage-2 models seem to matter more than the choices of stage-1 models as algorithms with the same stage-2 algorithms share analogous behavior albeit different stage-1 algorithms. Random Forests provided nontrivial improvements to parametric regressions with a lower MSE.

**Table 2.** MSE, relative MSE, R^2^, and Relative Efficiency (RE) for algorithms in MEPS analysis

| Algorithm | MSE<br>($10^9$) | Relative<br>MSE | $R^2$ | RE |
| --- | --- | --- | --- | --- |
| Two-stage Super Learner | 2.176 | 1.000 | 0.149 | 1.000 |
| Discrete Super Learner | 2.192 | 1.007 | 0.143 | 0.958 |
| S1: RF + S2: RF | 2.192 | 1.007 | 0.143 | 0.958 |
| S1: GLM + S2: RF | 2.193 | 1.008 | 0.143 | 0.956 |
| S1: Lasso + S2: RF | 2.194 | 1.008 | 0.142 | 0.953 |
| One-stage Super Learner | 2.226 | 1.023 | 0.130 | 0.869 |
| RF | 2.239 | 1.029 | 0.125 | 0.835 |
| Zero-inflated Poisson (ZIP) | 2.257 | 1.037 | 0.118 | 0.788 |
| S1: GLM + S2: Lasso (OLS) | 2.260 | 1.039 | 0.117 | 0.780 |
| S1: Lasso + S2: Lasso (OLS) | 2.260 | 1.039 | 0.117 | 0.780 |
| S1: RF + S2: Lasso (OLS) | 2.261 | 1.039 | 0.116 | 0.778 |
| OLS | 2.264 | 1.040 | 0.115 | 0.770 |
| Lasso (OLS) | 2.265 | 1.041 | 0.115 | 0.767 |
| S1: RF + S2: GLM-Gamma-Log | 2.267 | 1.042 | 0.114 | 0.762 |
| S1: GLM + S2: GLM-Gamma-Log | 2.268 | 1.042 | 0.114 | 0.759 |
| Zero-inflated Negative Binomial (ZINB) | 2.269 | 1.043 | 0.113 | 0.757 |
| S1: Lasso + S2: GLM-Gamma-Log | 2.270 | 1.043 | 0.113 | 0.754 |
| S1: RF + S2: Quantile regression | 2.271 | 1.044 | 0.112 | 0.752 |
| S1: RF + S2: GLM-Gamma-Identity | 2.272 | 1.044 | 0.112 | 0.749 |
| S1: GLM + S2: Quantile regression | 2.273 | 1.045 | 0.112 | 0.746 |
| S1: Lasso + S2: Quantile regression | 2.273 | 1.045 | 0.112 | 0.746 |
| S1: GLM + S2: GLM-Gamma-Identity | 2.275 | 1.045 | 0.111 | 0.741 |
| S1: Lasso + S2: GLM-Gamma-Identity | 2.275 | 1.045 | 0.111 | 0.741 |
| Tobit | 2.283 | 1.049 | 0.108 | 0.720 |
| Tweedie | 2.298 | 1.056 | 0.102 | 0.681 |
| S1: RF + S2: Adaptive hazard | 2.306 | 1.060 | 0.099 | 0.660 |
| S1: GLM + S2: Adaptive hazard | 2.311 | 1.062 | 0.097 | 0.647 |
| S1: Lasso + S2: Adaptive hazard | 2.311 | 1.062 | 0.097 | 0.647 |
| S1: RF + S2: Cox hazard | 2.355 | 1.082 | 0.080 | 0.532 |
| S1: GLM + S2: Cox hazard | 2.359 | 1.084 | 0.078 | 0.521 |
| S1: Lasso + S2: Cox hazard | 2.359 | 1.084 | 0.078 | 0.521 |
| S1: RF + S2: AFT (generalized Gamma) | 2.433 | 1.118 | 0.049 | 0.328 |
| S1: Lasso + S2: AFT (generalized Gamma) | 2.436 | 1.119 | 0.048 | 0.320 |
| S1: GLM + S2: AFT (generalized Gamma) | 2.436 | 1.119 | 0.048 | 0.320 |
| S1: RF + S2: Log OLS-smearing | 2.489 | 1.144 | 0.027 | 0.181 |
| S1: RF + S2: Adaptive GLM | 2.491 | 1.145 | 0.026 | 0.176 |
| S1: Lasso + S2: Log OLS-smearing | 2.492 | 1.145 | 0.026 | 0.174 |
| S1: Lasso + S2: Adaptive GLM | 2.493 | 1.146 | 0.026 | 0.171 |
| S1: GLM + S2: Log OLS-smearing | 2.494 | 1.146 | 0.025 | 0.168 |
| S1: GLM + S2: Adaptive GLM | 2.495 | 1.147 | 0.025 | 0.166 |
| OLS intercept only (mean) | 2.593 | 1.192 | -0.014 | -0.090 |

Algorithms are presented in ascending order based on MSE. S1 refers to stage-1 and S2 refers to stage-2. RF refers to Random Forest. GLM in S1 refers to logistic regression and Lasso in S1 refers to logistic Lasso regression. GLM-Gamma-Log refers to GLM with Gamma distribution and Log link function. Log-OLS smearing refers to logarithmic OLS with smearing retransformation. Lower MSE and relative MSE indicate better performance. Greater $R^2$ and RE indicates better performance.

#### BOLD

Distributions of four spine-related RVUs were all highly skewed with heavy upper tails but varied in both scales and levels of zero-inflation (Appendix-Table 8, Appendix-Figure 3). Spine-related RVUs had the lowest zero-inflation (5%) while spine-related injection RVUs had the greatest zero-inflation (91%).

In these data, we again found that two-stage super learner was the best performing algorithm under all different zero-inflation levels, although improvements over the discrete learner (average MSE improvement: 0.6%), one-stage super learner (average MSE improvement: 1.0%) and the best single algorithm (average MSE improvement: 0.8%) were more modest than in the MEPS data (Table 3). Performances of single algorithms changed dramatically under different zero-inflations levels (Appendix-E). For example, the random forest had strong performances for modeling spine-related RVUs (5% zero) and spine-related imaging RVUs (55% zero), but poor performances for modeling spine-related physical therapy RVUs (85% zero) and spine-related injection RVUs (91% zero). One-stage super learner and single-stage algorithms showed good performances when zero-inflation was low, as illustrated by the fact that the one-stage super learner was the second best-performing algorithm, and single-stage Random Forest is the best single algorithm in modeling spine-related RVUs.

However, these models did not perform as well for modeling spine-related injection RVUs, in which zero inflation is greater.

## Discussion

The novel two-stage super learner algorithm provided robust performance across a wide range of situations using both simulated and real data. We also found that the one-stage super learner performed well, and in some cases when zero inflation was small, even better than the two-stage super learner. We found that some individual algorithms had a good performance in certain scenarios, but poor performance in others. Since an analyst is unlikely to know, a priori, what the best performing single algorithm will be, we believe that super learning provides a useful general approach for estimating healthcare expenditures. In other words, super learning eliminates the need for a-priori knowledge as to which single algorithm might perform best in a given application--we found that it performed well in each scenario that we studied.

Our results are consistent with and extend prior research regarding the estimation of healthcare expenditures. First, we found that many commonly used methods do perform well in particular situations. However, we did not find a single algorithm that performed well in all situations. A major contribution of this study is that the novel two-stage super learning framework worked well across a wide variety of simulated and real-world datasets. Next, we found that the one-stage super learner also generally performed well. Though one-stage super learner is commonly used for other healthcare outcomes [27, 39, 40], its use is uncommon when working with healthcare expenditure data. We found that it often performs well and may deserve wider use.

Finally, the findings of this study help distinguish situations when one may want to use one-stage versus two-stage super learner approaches. When zero-inflation is low, the one-stage super learner may be a reasonable strategy. However, in the presence of higher zero-inflation, using a two-stage super learner is likely to lead to better performance.

This study suggests several directions for future work. First, we have focused on optimizing a super learner with respect to mean squared error. We believe this is an appropriate scientific goal in many situations and in particular, in situations where super learning is used as an intermediate step in drawing inference about the average causal effect. However, for some pure prediction tasks, mean squared error may not be the most appealing measure to gauge predictions since it is very sensitive to outliers. In these settings, it may be interesting to consider super learners constructed using alternative loss functions that are more robust to large costs. Second, our approach is focused on the prediction of the overall cost. In some circumstances, the scientific question may motivate us to simultaneously consider the prediction of having any cost (i.e., the stage-1 model itself is interesting) *as well as* the cost itself if some costs are present. For such problems, an alternative approach to super learning may be more useful, wherein one develops a super learner for each component of the model. In such an approach, each of the two super learners would be optimized towards a particular goal, as opposed to here where a single super learner is fit that is optimized towards the overall goal of predicting costs.

The findings of this study should be considered in light of several limitations. First, as with all simulation studies, we could not consider all possible data generation scenarios. While we picked a robust set of scenarios that likely reflect many real-life situations, we do not know if the results we observed generalize to other data generation processes that we did not examine. Similarly, as we analyzed only two empirical data sets, and the results may not generalize to other settings. In future work, we hope to examine more diverse scenarios to further inform practitioners in the application of our methods. An additional limitation is that we focused on super learning for the sake of prediction. A natural extension would be to study the potential benefits of two-stage super learning in the context of estimating the causal effect of an intervention on a cost outcome.

## Conclusions

In this study, we introduce a novel two-stage super learner approach that is well suited to many of the common problems encountered when analyzing healthcare expenditure data. Further, we provide a more in-depth study of the application of one-stage super learner to the healthcare expenditure context. We find that the two-stage super learner provides robust performance across a wide variety of data generating processes and that one-stage super learner also performs well, especially when there is little zero inflation. Given the notorious challenges that healthcare expenditures data present to the analyst, we think that super learner approaches will be a useful tool to advance health services research.

## Repository

The data and code used to train the two-stage super learner, one-stage super learner and individual algorithms can be found at https://github.com/wuziyueemory/Two-stage-SuperLearner. Introductions to simulations and real-data analysis are provided on the above GitHub page.

## Data Availability

The data used in simulation studies of this work are from the data generating process with code openly available at https://github.com/wuziyueemory/Two-stage-SuperLearner/blob/master/Simulation/createData.R.

The data used in MEPS empirical analysis is 3rd party data from the 2016-2017 Medical Expenditure Panel Survey (MEPS) database. This 3rd party data is free online (see https://www.meps.ahrq.gov/mepsweb/data_stats/download_data_files_detail.jsp?cboPufNumber=HC-202).

The data used in the BOLD empirical analysis is 3rd party data from The back pain outcomes using longitudinal data (BOLD) registry. The data could be accessed through https://github.com/wuziyueemory/Two-stage-SuperLearner/blob/master/BOLD%20data%20analysis/bold%20data.csv.

https://www.meps.ahrq.gov/mepsweb/data_stats/download_data_files_detail.jsp?cboPufNumber=HC-202

https://github.com/wuziyueemory/Two-stage-SuperLearner/blob/master/Simulation/createData.R

https://github.com/wuziyueemory/Two-stage-SuperLearner/blob/master/BOLD%20data%20analysis/bold%20data.csv

## Acknowledgments

Research reported in this publication was supported by the University of Washington Clinical Learning, Evidence And Research (CLEAR) Center for Musculoskeletal Research. The CLEAR Center is supported by the National Institute of Arthritis and Musculoskeletal and Skin Diseases (NIAMS) of the National Institutes of Health under Award Number P30AR072572. The content is solely the responsibility of the authors and does not necessarily represent the official views of the National Institutes of Health. We thank Jerry Jarvik, Sandra Johnston, and Laurie Gold for helpful comments and providing data on the BOLD analysis in this study.

## Appendix

### Technical appendix

#### A. Additional modifications of two-stage super learner

We consider two modifications on (i) the weights calculation and (ii) the cross-validation scheme to improve the two-stage super learner on predicting healthcare costs. For continuous outcome, a quadratic programming algorithm designed by Goldfarb and Idnani was generally applied to calculate the best convex combination of weights that minimize the squared error loss. However, the heavy upper tails in healthcare expenditure would result in excessive huge numbers in the matrix and vector of a quadratic function to be minimized. This consequently induces overflow errors and causes the quadratic programming constraints inconsistent. As a modification, we proposed a scaling scheme that divides the quadratic function by a large constant to shrink the huge matrix and vector in the quadratic function. The scaling would not affect the results given the raw quadratic function is just a multiple of the scaled quadratic function. Alternatively, we consider a modification to the cross-validation scheme in the standard super learning procedure. The standard V-fold cross-validation allocates subjects randomly to each block. However, this random allocation could result in all observed subjects with zero costs or large costs in the same block of data. We might expect a better finite sample evaluation of how well methods fit zero as well as large costs by evenly splitting the zero and large costs amongst the blocks. Considering the minimum of our data is zero, we propose a snake-like assignment of subjects to each block. That is, the V lowest ordered costs are assigned to blocks 1 through V, then the next V lowest ordered costs are assigned reversely to blocks V through 1, respectively. This process is repeated until all subjects have been assigned a block. By splitting up the zero and largest costs, we ensure that every time we fit the candidate methods to V-1 of the blocks, there will be an adequate number of subjects with zero and large costs in the held-out block.

#### B. Regression formula for zero-inflation & non-zero cost distribution

Costs were simulated using a two-stage procedure to allow for different zero-inflation levels and non-zero cost distributions. In stage-1, we drew a random variable *Z* from a Bernoulli distribution with the probability of zero determined by a logistic model:=

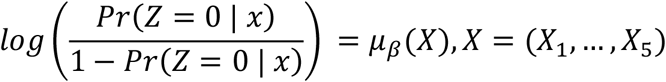

The regression formula *μ*_*β*_(*X*) was mainly controlled by zero-inflation percentage and data complexity (Table 1). If *Z* = 0, then we set costs equal to zero. If *Z* = 1, we drew the observed cost from one of the four different non-zero cost distributions (Log-normal, Gamma, Tweedie, Mixture) with the corresponding parameter mean (Log-normal, Tweedie) and shape (Gamma) generated by another regression formula *μ*_*λ*_(*X*), *X* = (*X*_1_, …, *X*_5_). The regression formula *μ*_*λ*_(*X*) was mainly controlled by data complexity (Table 2).

**Table 1:**
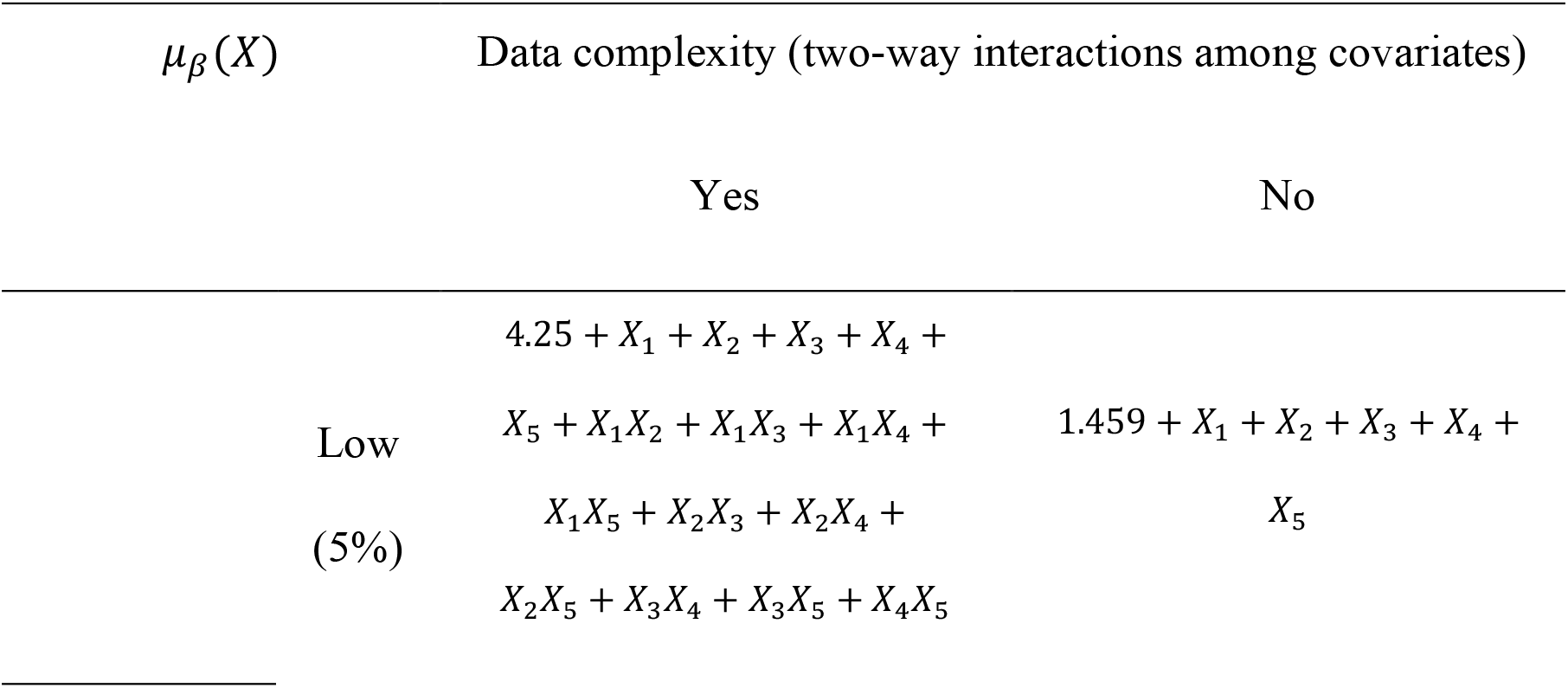
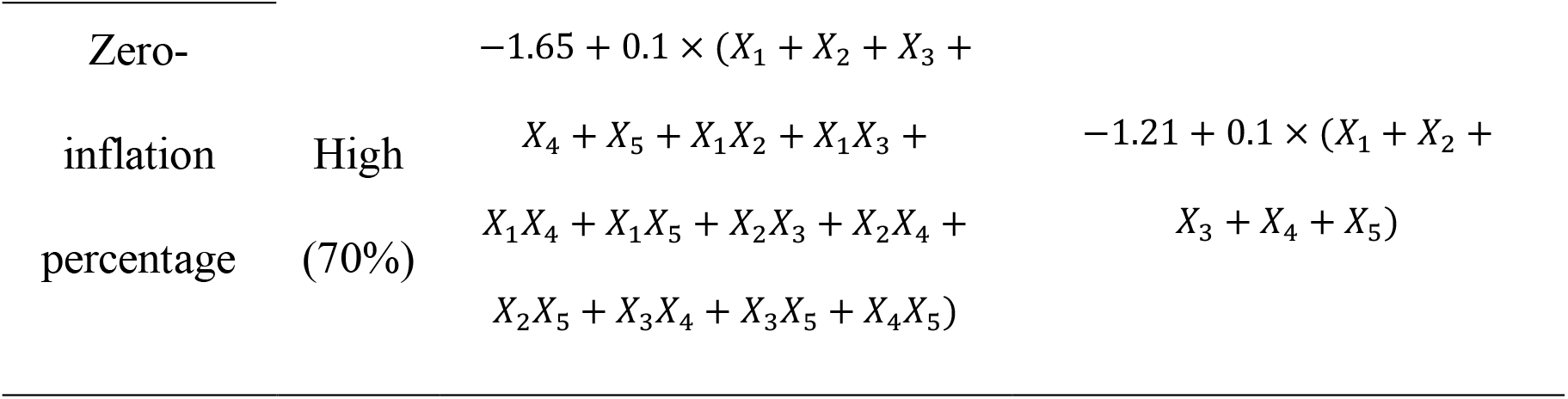
Stage-1 regression formulas *μ*_*β*_(*X*) under different zero-inflation levels and data complexities

| $\mu_\beta(X)$ | Data complexity (two-way interactions among covariates) | |
| --- | --- | --- |
|  | Yes | No |
| Low<br>(5%) | 4.25 + $X_1$ + $X_2$ + $X_3$ + $X_4$ + | |
| | $X_5$ + $X_1X_2$ + $X_1X_3$ + $X_1X_4$ + | 1.459 + $X_1$ + $X_2$ + $X_3$ + $X_4$ + |
| | $X_1X_5$ + $X_2X_3$ + $X_2X_4$ + | $X_5$ |
| | $X_2X_5$ + $X_3X_4$ + $X_3X_5$ + $X_4X_5$ | |

|  |  |  |  |
| --- | --- | --- | --- |
| Zero- | | $-1.65 + 0.1 \times (X_1 + X_2 + X_3 +$ | |
| inflation | High | $X_4 + X_5 + X_1X_2 + X_1X_3 +$ | $-1.21 + 0.1 \times (X_1 + X_2 +$ |
| percentage | (70%) | $X_1X_4 + X_1X_5 + X_2X_3 + X_2X_4 +$ | $X_3 + X_4 + X_5)$ |
| | | $X_2X_5 + X_3X_4 + X_3X_5 + X_4X_5)$ | |

**Table 2:** Stage-2 regression formulas *μ*_*λ*_(*X*) under different data complexities

| | | $\mu_\lambda(X)$ |
| --- | --- | --- |
| Data | Yes | $1 + 0.3 \times X_1 - X_2 + 0.2 \times X_3 - 0.15 \times X_4 + 0.1 \times X_5 - 0.01 \times$ |
| complexity | | $X_1X_2 +$ |
| (two-way | | $0.03 \times X_1X_3 - 0.1 \times X_2X_3 + 0.02 \times X_3X_4 - 0.04 \times X_4X_5$ |
| interactions | No | $0.1 + 0.5 \times X_1 - 1.5 \times X_2 + 0.4 \times X_3 - 0.3 \times X_4 - 0.2 \times X_5$ |
| among |  |  |
| covariates) |  |  |

#### C. Data generating process - distribution of non-zero costs

We generate non-zero costs from one of the four distributions (Log-normal, Gamma, Tweedie, Mixture). The mean parameter *μ* for the natural logarithm and Tweedie distributions and shape parameter *β* for the Gamma distribution was controlled by the regression formula *μ*_*λ*_(*X*), *X* = (*X*_1_, …, *X*_5_) described in table 2 above.

#### Lognormal Distribution

The log-normal model for positive costs can be written as *In* (*y*) = *μ*_*λ*_(*X*) + *ε*, where *X* = (*X*_1_, …, *X*_5_), *ε ∼ N*(0, *σ*^2^), *E*(*X*^*T*^*ε*) = 0. The conditional expectation of *y* given *y* > 0 can thus be written as *E*(*y* | *x*) = *exp* (*μ*_*λ*_(*X*) + 0.5*σ*^2^). We set *σ*^2^ = 0.3.

#### Gamma Distribution

The Gamma distribution has PDF 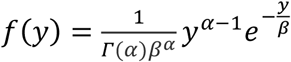 where *β* is the shape parameter and *α* is the scale parameter. The mean is *αβ*. The shape parameter was set to *β* = *exp* (*μ*_*λ*_(*X*)) and the scale parameter was set to *α* = 1.3.

#### Tweedie Distribution

Tweedie distributions are defined as a subfamily of exponential dispersion models (ED) with a special mean-variance relationship. A random variable *Y* is Tweedie distributed *Tw*_*p*_(*μ, σ*^2^), if its mean *μ* = *E*(*Y*) and *var*(*Y*) = *σ*^2^*μ*^*p*^, where *σ*^2^ is the dispersion parameter and *p ∈ R* is the power parameter. The PDF for the Tweedie family is complex and cannot be expressed in closed form, but the Tweedie family includes common distributions like Normal (*p* = 0), Poisson (*p* = 1), and Gamma (*p* = 2). In the simulation, the mean was set to *μ* = *μ*_*λ*_(*X*), while the scale and power were set to *σ*^2^ = 1.8, and *p* = 1.5, respectively.

#### Mixture distribution

The mixture distribution is a mixture of log-normal and gamma distribution. Specifically, it was generated by first drawing a binary random variable with a pre-specified probability, *G ∼ BernouIIi*(*p*). We subsequently generated a gamma distribution if *G* ≤ *p*, or a log-normal distribution if *G* > *p*. In the simulation, the probability was set to *p* = 0.5. We wanted to evaluate the two-stage super learner in situations where a simple parametric model does not capture the true distribution, as we expect to be the case in practice.

#### D. Details of covariates used in the BOLD study

In this study, patient self-reported questionnaire responses were used to predict future expenditures (measured by RVUs) in several categories. Specifically, the covariates include the following measures from patient self-reported questionnaires collected at baseline: (1) Socio-demographics (age, sex, race, ethnicity, education, employment status, etc.); (2) Pain-related characteristics (back/leg pain duration, back/leg pain intensity, modified Roland-Morris Disability Questionnaire, Brief Pain Inventory Activity Interference Scale); (3) PHQ-4 measure of anxiety and depressive symptoms; (4) European Quality of Life 5 Dimension (EQ5D) index and Visual Analog Scale; (5) Number of falls; and (6) Recovery expectation. Besides, we also include the Quan comorbidity score, baseline diagnosis, and total RVUs at one year before index visit from EHR as covariates.

#### E. Extended results of BOLD analyses

Individual algorithms that perform well in one setting will not necessarily perform well in other settings. The best individual algorithms in modeling 4 spine-related RVUs changed dramatically. One-stage super learner worked better in modeling outcomes with low to medium zero-inflation, with performance ranked 2^*t*h^ in modeling spine-related RVUs (5% zero) and 6^*t*h^ in modeling spine-related imaging RVUs (55% zero). One-stage individual algorithms had good behavior likewise when zero-mass were not serious, especially in low zero-inflation situations where the one-stage Random Forest beat all two-stage individual algorithms in modeling spine-related RVUs (5% zero). Baseline socio-demographic and patient-reported outcomes may have signals for predicting various spine-related RVUs in a year after index, with a cross-validated *R*^2^ ranging from 6.2% to 27.8% for the two-stage super learner. The performance was especially great for modeling spine-related physical therapy RVUs. MSE varied significantly in modeling different spine-related RVUs, partly due to the difference in terms of the scale of RVUs. The results also suggested that there may exist redundancy in the covariates as Lasso was always among the best-performed algorithms in modeling spine-related RVUs with different zero-inflations.

## Tables

**Table 3:** Algorithms for Monte Carlo Simulations and MEPS analysis

| Super Learner | Stage | Method |
| --- | --- | --- |
| Two-stage | 1 | GLM (logistic regression) |
|  | 1 | Lasso (logistic regression) |
|  | 1 | Random Forest |
|  | 2 | GLM (log link, Gamma family) |
|  | 2 | GLM (identity link, Gamma family) |
|  | 2 | Log OLS + smearing |
|  | 2 | Lasso (OLS) |
|  | 2 | Adaptive GLM |
|  | 2 | AFT (generalized Gamma) |
|  | 2 | Cox hazard |
|  | 2 | Adaptive hazard |
|  | 2 | Quantile regression |
|  | 2 | Random forest |
| Standard (one-stage) |  | GLM intercept only (mean) |
|  |  | OLS |
|  |  | Lasso (OLS) |
|  |  | Zero-inflated Poisson |
|  |  | Zero-inflated Negative Binomial |
|  |  | Tobit |
|  |  | Tweedie |
|  |  | Random Forest |

**Table 4:** MSE, relative MSE, R^2^, and Relative Efficiency (RE) for all algorithms in the Monte Carlo simulations

| Algorithm | MSE<br>($10^8$) | Relative<br>MSE | $R^2$ | RE |
| --- | --- | --- | --- | --- |
| Two-stage Super Learner | 3.239 | 1.000 | 0.659 | 1.000 |
| Discrete Super Learner | 3.301 | 1.019 | 0.646 | 0.980 |
| One-stage Super Learner | 3.374 | 1.042 | 0.631 | 0.958 |
| S1: Lasso + S2: GLM-Gamma-Log | 3.563 | 1.100 | 0.610 | 0.926 |
| S1: Lasso + S2: Quantile regression | 3.567 | 1.101 | 0.609 | 0.924 |
| S1: Lasso + S2: Log OLS-smearing | 3.570 | 1.102 | 0.609 | 0.924 |
| Zero-inflated Negative Binomial (ZINB) | 3.582 | 1.106 | 0.608 | 0.923 |
| S1: GLM + S2: GLM-Gamma-Log | 3.583 | 1.106 | 0.608 | 0.923 |
| S1: GLM + S2: Quantile regression | 3.589 | 1.108 | 0.607 | 0.921 |
| S1: GLM + S2: Log OLS-smearing | 3.590 | 1.108 | 0.607 | 0.921 |
| Zero-inflated Poisson (ZIP) | 3.601 | 1.112 | 0.606 | 0.920 |
| S1: Lasso + S2: Adaptive GLM | 3.784 | 1.168 | 0.593 | 0.900 |
| S1: GLM + S2: Adaptive GLM | 3.803 | 1.174 | 0.591 | 0.897 |
| S1: RF + S2: GLM-Gamma-Log | 4.381 | 1.353 | 0.550 | 0.835 |
| S1: RF + S2: Quantile regression | 4.385 | 1.354 | 0.548 | 0.832 |
| S1: RF + S2: Log OLS-smearing | 4.390 | 1.355 | 0.545 | 0.827 |
| S1: RF + S2: Adaptive GLM | 4.628 | 1.429 | 0.533 | 0.809 |
| S1: GLM + S2: RF | 4.898 | 1.512 | 0.523 | 0.794 |
| S1: Lasso + S2: RF | 4.900 | 1.513 | 0.522 | 0.792 |
| RF | 5.304 | 1.638 | 0.496 | 0.753 |
| S1: RF + S2: RF | 5.506 | 1.700 | 0.486 | 0.737 |
| S1: Lasso + S2: AFT (generalized Gamma) | 6.321 | 1.952 | 0.445 | 0.675 |
| S1: GLM + S2: AFT (generalized Gamma) | 6.339 | 1.957 | 0.443 | 0.672 |
| S1: RF + S2: AFT (generalized Gamma) | 7.049 | 2.176 | 0.432 | 0.656 |
| Tobit | 9.165 | 2.830 | 0.366 | 0.555 |
| S1: Lasso + S2: Lasso (OLS) | 9.295 | 2.870 | 0.351 | 0.533 |
| S1: GLM + S2: Lasso (OLS) | 9.313 | 2.875 | 0.348 | 0.528 |
| Lasso (OLS) | 9.677 | 2.988 | 0.310 | 0.470 |
| OLS | 9.703 | 2.996 | 0.304 | 0.461 |
| S1: RF + S2: Lasso (OLS) | 9.907 | 3.059 | 0.298 | 0.452 |
| S1: Lasso + S2: Adaptive hazard | 9.920 | 3.063 | 0.282 | 0.428 |
| S1: GLM + S2: Adaptive hazard | 9.933 | 3.067 | 0.281 | 0.426 |
| S1: RF + S2: Adaptive hazard | 10.226 | 3.157 | 0.257 | 0.390 |
| Tweedie | 11.273 | 3.480 | 0.245 | 0.372 |
| S1: GLM + S2: Cox hazard | 12.341 | 3.810 | 0.236 | 0.358 |
| S1: Lasso + S2: Cox hazard | 12.353 | 3.814 | 0.235 | 0.357 |
| S1: RF + S2: Cox hazard | 12.820 | 3.958 | 0.225 | 0.341 |
| S1: GLM + S2: GLM-Gamma-Identity | 13.249 | 4.090 | 0.213 | 0.323 |
| S1: Lasso + S2: GLM-Gamma-Identity | 13.263 | 4.095 | 0.211 | 0.320 |
| S1: RF + S2: GLM-Gamma-Identity | 13.717 | 4.235 | 0.167 | 0.253 |
| OLS intercept only (mean) | 17.512 | 5.406 | -0.002 | -0.003 |

Lower MSE indicates better performance. Algorithms are presented in ascending order according to the overall mean MSE. S1 refers to stage-1 and S2 refers to stage-2. GLM in S1 refers to logistic regression and Lasso in S1 refers to logistic Lasso regression. RF refers to Random Forest. GLM-Gamma-Log refers to GLM with Gamma distribution and Log link function.

**Table 5:**
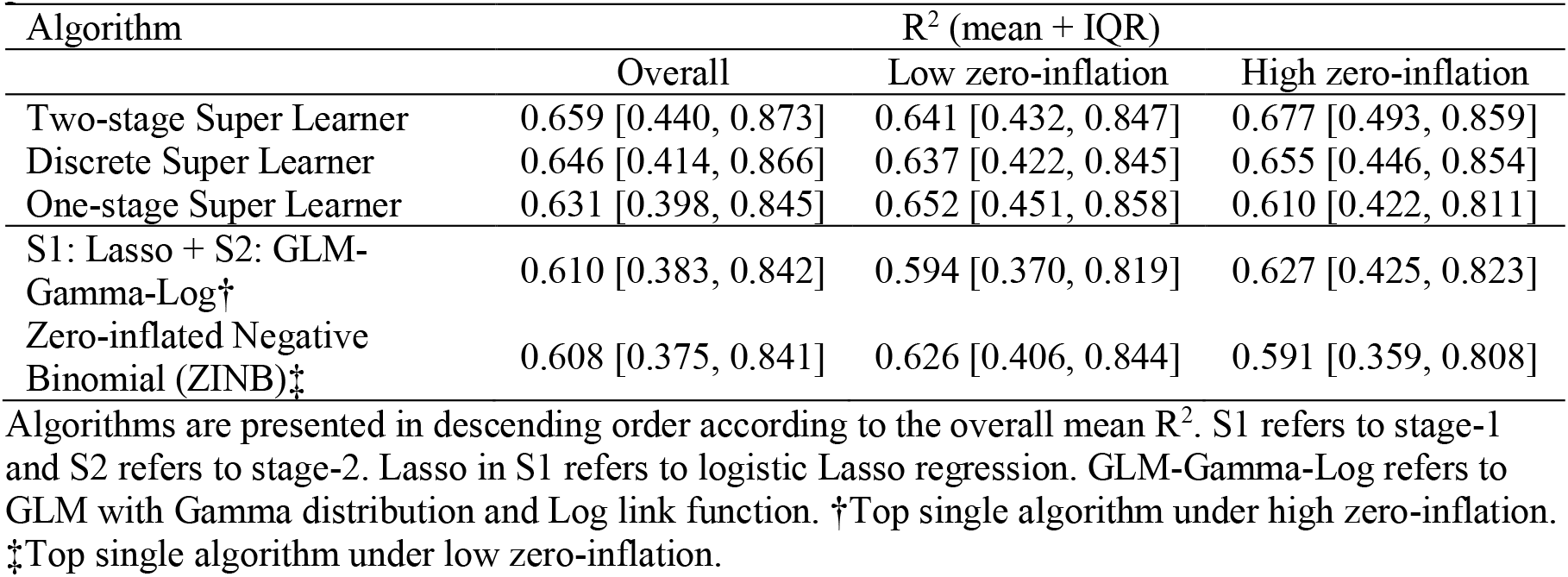
Average and IQR of R^2^ for selected algorithms across 32 data generating processes

**Table 6:** Summary statistics for annual healthcare expenditures of MEPS data

| Measures | 2016 MEPS<br>N=10925 | 2017 MEPS<br>N=10815 |
| --- | --- | --- |
| 0.05 Percentile | 0.0 | 0.0 |
| 0.25 Percentile | 157.0 | 130.0 |
| 0.50 Percentile | 1076.0 | 1156.0 |
| 0.75 Percentile | 4521.0 | 4902.0 |
| 0.95 Percentile | 24277.8 | 28189.3 |
| Mean | 5368.4 | 6099.7 |
| SD | 13815.1 | 15992.3 |
| Skewness | 7.2 | 6.7 |
| Zero percentage | 17.8 | 19.6 |

The distribution of annual healthcare expenditures is highly skewed with a large mass at zero. The skewness is 7.2 and 6.7 for 2016 & 2017 MEPS data, respectively (compare to 0 for symmetric data). Almost 20% of observations have zero expenditures and in a very small fraction of observations, 2% to be precise, had expenditures over \$50,000.

**Table 7:**
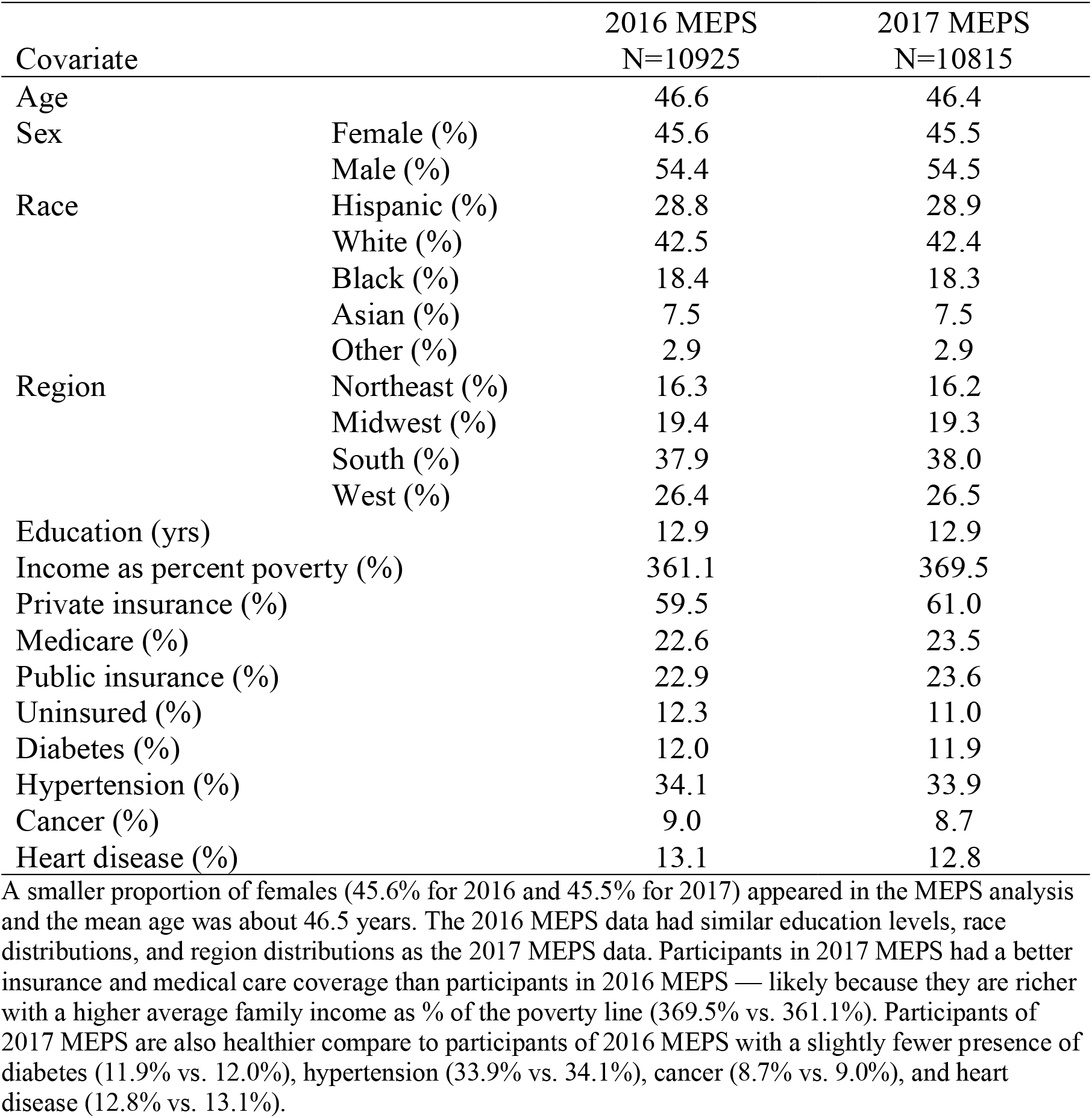
Descriptive statistics of baseline covariates in MEPS data

| Covariate |  | 2016 MEPS<br>N=10925 | 2017 MEPS<br>N=10815 |
| --- | --- | --- | --- |
| Age |  | 46.6 | 46.4 |
| Sex | Female (%) | 45.6 | 45.5 |
|  | Male (%) | 54.4 | 54.5 |
| Race | Hispanic (%) | 28.8 | 28.9 |
|  | White (%) | 42.5 | 42.4 |
|  | Black (%) | 18.4 | 18.3 |
|  | Asian (%) | 7.5 | 7.5 |
|  | Other (%) | 2.9 | 2.9 |
| Region | Northeast (%) | 16.3 | 16.2 |
|  | Midwest (%) | 19.4 | 19.3 |
|  | South (%) | 37.9 | 38.0 |
|  | West (%) | 26.4 | 26.5 |
| Education (yrs) |  | 12.9 | 12.9 |
| Income as percent poverty (%) |  | 361.1 | 369.5 |
| Private insurance (%) |  | 59.5 | 61.0 |
| Medicare (%) |  | 22.6 | 23.5 |
| Public insurance (%) |  | 22.9 | 23.6 |
| Uninsured (%) |  | 12.3 | 11.0 |
| Diabetes (%) |  | 12.0 | 11.9 |
| Hypertension (%) |  | 34.1 | 33.9 |
| Cancer (%) |  | 9.0 | 8.7 |
| Heart disease (%) |  | 13.1 | 12.8 |

A smaller proportion of females (45.6% for 2016 and 45.5% for 2017) appeared in the MEPS analysis and the mean age was about 46.5 years. The 2016 MEPS data had similar education levels, race distributions, and region distributions as the 2017 MEPS data. Participants in 2017 MEPS had a better insurance and medical care coverage than participants in 2016 MEPS — likely because they are richer with a higher average family income as % of the poverty line (369.5% vs. 361.1%). Participants of 2017 MEPS are also healthier compare to participants of 2016 MEPS with a slightly fewer presence of diabetes (11.9% vs. 12.0%), hypertension (33.9% vs. 34.1%), cancer (8.7% vs. 9.0%), and heart disease (12.8% vs. 13.1%).

**Table 8:** Summary statistics for 4 spine-related RVUs of BOLD data

| Measures | Spine-related<br>RVUS | Spine-related<br>physical therapy<br>RVU | Spine-related<br>injection RVU | Spine-related<br>imaging RVU |
| --- | --- | --- | --- | --- |
| 0.05 Percentile | 0.10 | 0.00 | 0.00 | 0.00 |
| 0.25 Percentile | 1.48 | 0.00 | 0.00 | 0.00 |
| 0.50 Percentile | 2.45 | 0.00 | 0.00 | 0.00 |
| 0.75 Percentile | 7.38 | 0.00 | 0.00 | 0.00 |
| 0.95 Percentile | 28.32 | 3.39 | 4.28 | 15.40 |
| Mean | 14.83 | 0.54 | 0.75 | 3.30 |
| SD | 80.60 | 1.85 | 4.20 | 7.69 |
| Skewness | 12.01 | 6.20 | 13.96 | 8.20 |
| Zero percentage | 5.00 | 85.35 | 91.31 | 55.22 |

Distributions of 4 spine-related RVUs are all highly skewed with heavy upper tails despite the difference in scale. The skewness are all above 6, compared to 0 of absolute symmetry. Conversely, different spine-related RVUs have different zero inflation, with spine-related RVU having the lowest zero-mass (5%), then comes the spine-related imaging RVU (55%), spine-related physical therapy RVU (85%), and spine-related injection RVU (91%).

**Table 9:**
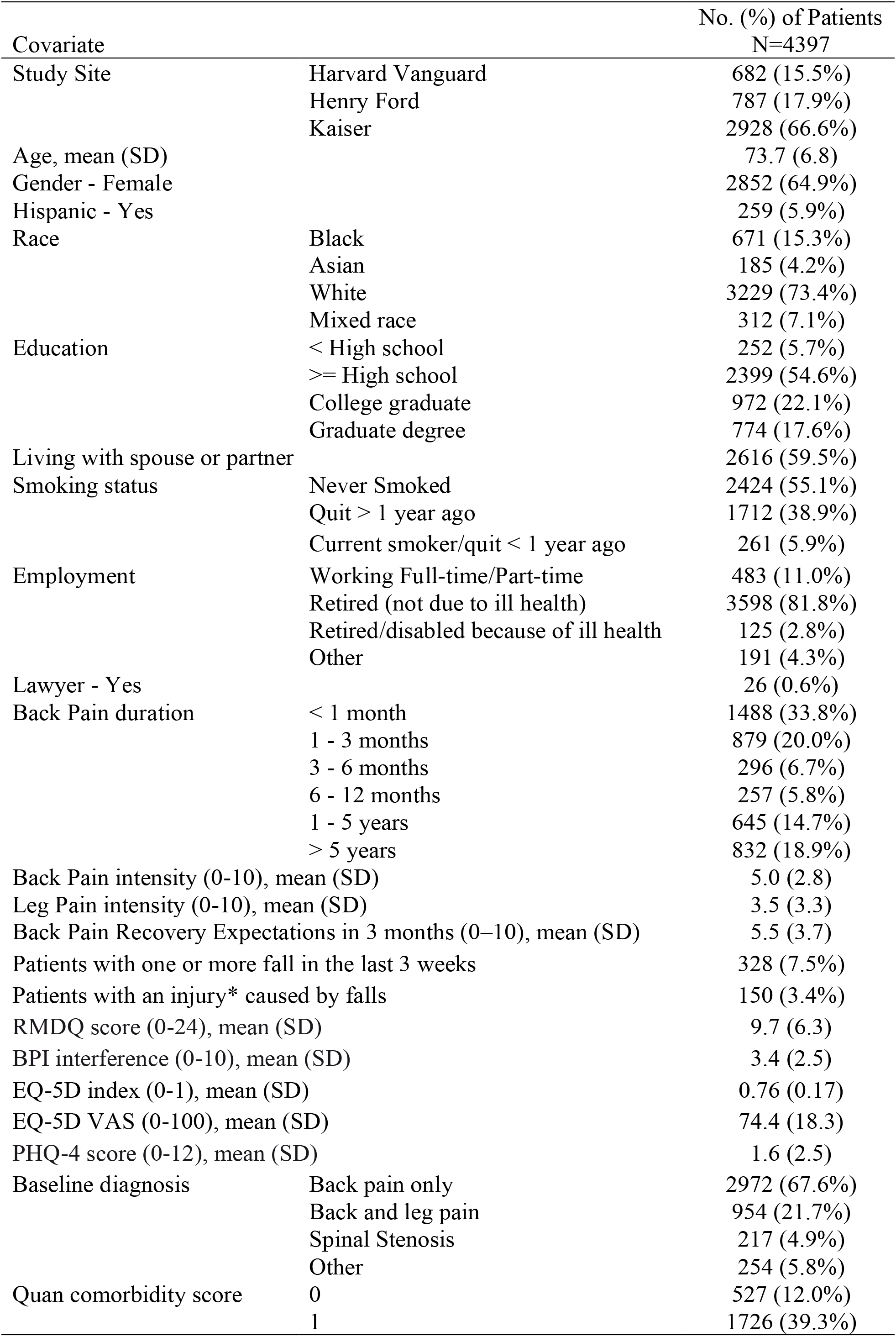
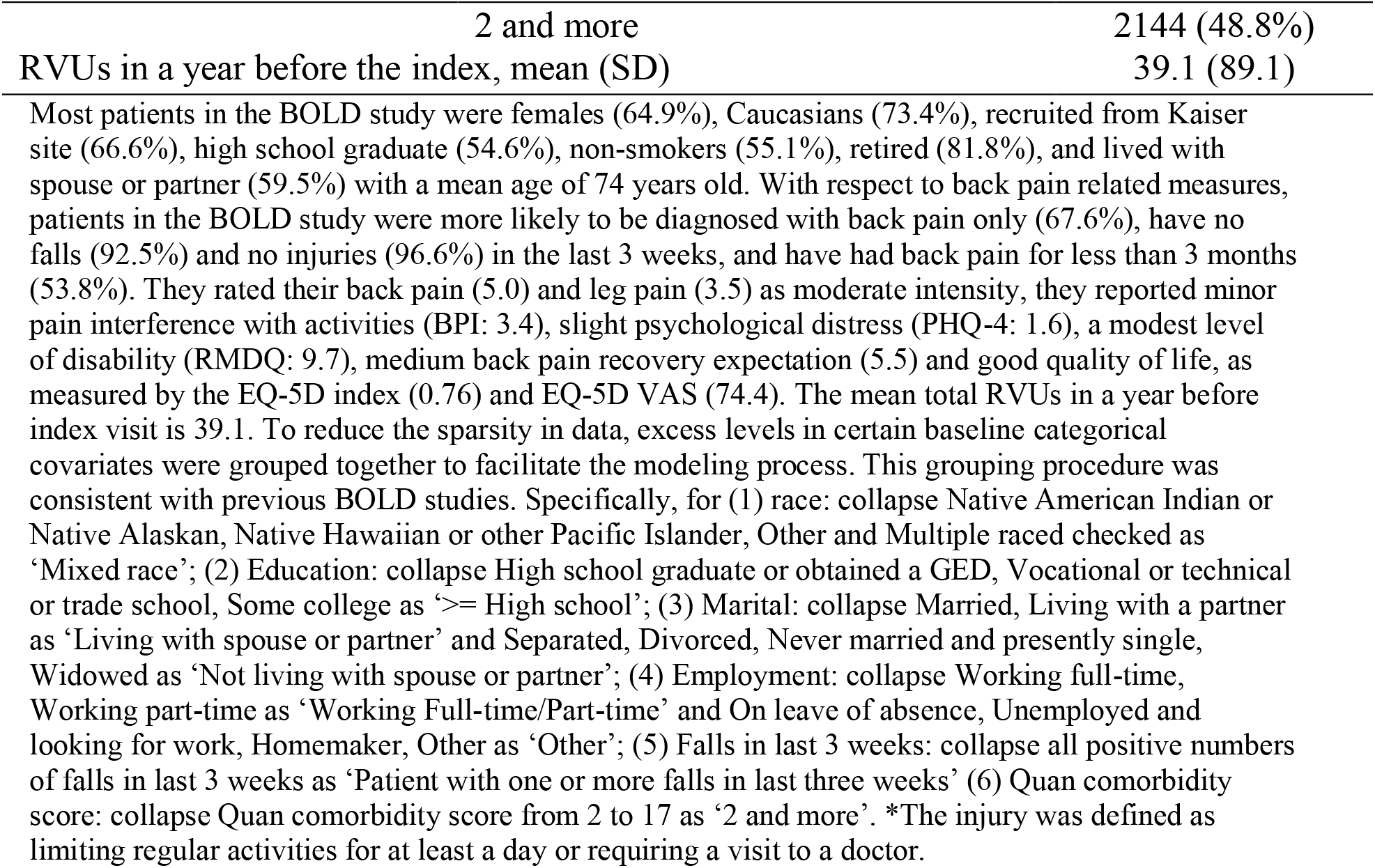
Descriptive statistics of baseline covariates in BOLD data

| Covariate |  | No. (%) of Patients<br>N=4397 |
| --- | --- | --- |
| Study Site | Harvard Vanguard | 682 (15.5%) |
|  | Henry Ford | 787 (17.9%) |
|  | Kaiser | 2928 (66.6%) |
| Age, mean (SD) |  | 73.7 (6.8) |
| Gender - Female |  | 2852 (64.9%) |
| Hispanic - Yes |  | 259 (5.9%) |
| Race | Black | 671 (15.3%) |
|  | Asian | 185 (4.2%) |
|  | White | 3229 (73.4%) |
|  | Mixed race | 312 (7.1%) |
| Education | < High school | 252 (5.7%) |
|  | >= High school | 2399 (54.6%) |
|  | College graduate | 972 (22.1%) |
|  | Graduate degree | 774 (17.6%) |
| Living with spouse or partner |  | 2616 (59.5%) |
| Smoking status | Never Smoked | 2424 (55.1%) |
|  | Quit > 1 year ago | 1712 (38.9%) |
|  | Current smoker/quit < 1 year ago | 261 (5.9%) |
| Employment | Working Full-time/Part-time | 483 (11.0%) |
|  | Retired (not due to ill health) | 3598 (81.8%) |
|  | Retired/disabled because of ill health | 125 (2.8%) |
|  | Other | 191 (4.3%) |
| Lawyer - Yes |  | 26 (0.6%) |
| Back Pain duration | < 1 month | 1488 (33.8%) |
|  | 1 - 3 months | 879 (20.0%) |
|  | 3 - 6 months | 296 (6.7%) |
|  | 6 - 12 months | 257 (5.8%) |
|  | 1 - 5 years | 645 (14.7%) |
|  | > 5 years | 832 (18.9%) |
| Back Pain intensity (0-10), mean (SD) |  | 5.0 (2.8) |
| Leg Pain intensity (0-10), mean (SD) |  | 3.5 (3.3) |
| Back Pain Recovery Expectations in 3 months (0–10), mean (SD) |  | 5.5 (3.7) |
| Patients with one or more fall in the last 3 weeks |  | 328 (7.5%) |
| Patients with an injury* caused by falls |  | 150 (3.4%) |
| RMDQ score (0-24), mean (SD) |  | 9.7 (6.3) |
| BPI interference (0-10), mean (SD) |  | 3.4 (2.5) |
| EQ-5D index (0-1), mean (SD) |  | 0.76 (0.17) |
| EQ-5D VAS (0-100), mean (SD) |  | 74.4 (18.3) |
| PHQ-4 score (0-12), mean (SD) |  | 1.6 (2.5) |
| Baseline diagnosis | Back pain only | 2972 (67.6%) |
|  | Back and leg pain | 954 (21.7%) |
|  | Spinal Stenosis | 217 (4.9%) |
|  | Other | 254 (5.8%) |
| Quan comorbidity score | 0 | 527 (12.0%) |
|  | 1 | 1726 (39.3%) |

|  |  |
| --- | --- |
| 2 and more | 2144 (48.8%) |
| RVUs in a year before the index, mean (SD) | 39.1 (89.1) |

**Table 10:** Candidate algorithms for BOLD analysis

| Super Learner | Stage | Method |
| --- | --- | --- |
| Two-stage | 1 | GLM (logistic regression) |
|  | 1 | Lasso (logistic regression) |
|  | 1 | CV-Random Forest |
|  | 2 | GLM (log link, Gamma family) |
|  | 2 | GLM (identity link, Gamma family) |
|  | 2 | Log OLS + smearing |
|  | 2 | Lasso (OLS) |
|  | 2 | Multivariate Adaptive Regression Spline (MARS) |
|  | 2 | Regression Tree |
|  | 2 | Bootstrap Aggregating (Bagging) |
|  | 2 | CV-Random Forest |
|  | 2 | Gradient Boosting Machine |
|  | 2 | Neural Network |
| Standard (one-stage) |  | OLS |
|  |  | Lasso (OLS) |
|  |  | Multivariate Adaptive Regression Spline (MARS) |
|  |  | Regression Tree |
|  |  | Bootstrap Aggregating (Bagging) |
|  |  | CV-Random Forest |
|  |  | Gradient Boosting Machine |
|  |  | Neural Network |

## References

1. Dario Gregori, Michele Petrinco, Simona Bo, Alessandro Desideri, Franco Merletti, and Eva Pagano. Regression models for analyzing costs and their determinants in health care: an introductory review. International Journal for Quality in Health Care, 23(3):331–341, 04 2011.

2. Willard G Manning and John Mullahy. Estimating log models: to transform or not to transform? Journal of Health Economics, 20(4):461–494, 2001.

3. A.M Jones. Models For Health Care. Health, Econometrics and Data Group (HEDG) Working Papers 10/01, HEDG, c/o Department of Economics, University of York, January 2010.

4. Marc L. Berk and Alan C. Monheit. The concentration of health care expenditures, revisited. Health Affairs, 20(2):9–18, 2001. PMID: 11260963.

5. Naihua Duan. Smearing estimate: A nonparametric retransformation method. Journal of the American Statistical Association, 78(383):605–610, 1983.

6. David K. Blough, Carolyn W. Madden, and Mark C. Hornbrook. Modeling risk using generalized linear models. Journal of Health Economics, 18(2):153–171, 1999.

7. Willard G. Manning, Anirban Basu, and John Mullahy. Generalized modeling approaches to risk adjustment of skewed outcomes data. Journal of Health Economics, 24(3):465–488, 2005.

8. D. R. Cox. Regression models and life-tables. Journal of the Royal Statistical Society. Series B (Methodological), 34(2):187–220, 1972.

9. Donna B. Gilleskie and Thomas A. Mroz. A flexible approach for estimating the effects of covariates on health expenditures. Journal of Health Economics, 23(2):391–418, 2004.

10. Anirban Basu and Paul J. Rathouz. Estimating marginal and incremental effects on health outcomes using flexible link and variance function models. Biostatistics, 6(1):93–109, 01 2005.

11. Huixia Judy Wang and Xiao-Hua Zhou. Estimation of the retransformed conditional mean in health care cost studies. Biometrika, 97(1):147–158, 03 2010.

12. John Mullahy. Much ado about two: reconsidering retransformation and the two-part model in health econometrics. Journal of Health Economics, 17(3):247–281, 1998.

13. John Cawley and Chad Meyerhoefer. The medical care costs of obesity: An instrumental variables approach. Journal of Health Economics, 31(1):219–230, 2012.

14. Eric A Finkelstein, Justin G Trogdon, Joel W Cohen, and William Dietz. Annual medical spending attributable to obesity: payer-and service-specific estimates. Health affairs (Project Hope), 28(5):w822—31, 2009.

15. Benjamin Lê Cook, Thomas G. McGuire, Kari Lock, and Alan M. Zaslavsky. Comparing methods of racial and ethnic disparities measurement across different settings of mental health care. Health Services Research, 45(3):825– 847, 2010.

16. Anirban Basu and Willard G. Manning. Issues for the next generation of health care cost analyses. Medical Care, 47(7):S109–S114, 2009.

17. Gareth James, Daniela Witten, Trevor Hastie, and Robert Tibshirani. An Introduction to Statistical Learning: with Applications in R. Springer, 2013.

18. Anna Zink and Sherri Rose. Fair regression for health care spending. Biometrics, 76(3):973–982, 2020.

19. Sherri Rose. A machine learning framework for plan payment risk adjustment. Health Services Research, 51(6):2358–2374, 2016.

20. Akritee Shrestha, Savannah Bergquist, Ellen Montz, and Sherri Rose. Mental health risk adjustment with clinical categories and machine learning. Health Services Research, 53(S1):3189–3206, 2018.

21. Mohammad Amin Morid, Kensaku Kawamoto, Travis Ault, Josette Dorius, and Samir Abdelrahman. Supervised learning methods for predicting healthcare costs: Systematic literature review and empirical evaluation. AMIA … Annual Symposium proceedings. AMIA Symposium, 2017:1312—1321, 2017.

22. David H. Wolpert. Stacked generalization. Neural Networks, 5(2):241–259, 1992.

23. Leo Breiman. Stacked regressions. Mach. Learn., 24(1):49–64, July 1996.

24. Michael Leblanc and Robert Tibshirani. Combining estimates in regression and classification. Journal of the American Statistical Association, 91(436):1641– 1650, 1996.

25. Mark J van der Laan, Eric C Polley, and Alan E Hubbard. Super learner. Statistical applications in genetics and molecular biology, 6: Article 25, 2007.

26. Ronald C. Kessler, Sherri Rose, Karestan C. Koenen, Elie G. Karam, Paul E. Stang, Dan J. Stein, Steven G. Heeringa, Eric D. Hill, Israel Liberzon, Katie A. McLaughlin, Samuel A. McLean, Beth E. Pennell, Maria Petukhova, Anthony J. Rosellini, Ayelet M. Ruscio, Victoria Shahly, Arieh Y. Shalev, Derrick Silove, Alan M. Zaslavsky, Matthias C. Angermeyer, Evelyn J. BrometJosé, Miguel Caldas de Almeida, Giovanni de Girolamo, Peter de Jonge, Koen Demyttenaere, Silvia E. Florescu, Oye Gureje, Josep Maria Haro, Hristo Hinkov, Norito Kawakami, Viviane Kovess-Masfety, Sing Lee, Maria Elena Medina-Mora, Samuel D. Murphy, Fernando Navarro-Mateu, Marina Piazza, Jose Posada-Villa, Kate Scott, Yolanda Torres, and Maria Carmen Viana. How well can post-traumatic stress disorder be predicted from pre-trauma risk factors? an exploratory study in the who world mental health surveys. World Psychiatry, 13(3):265–274, 2014.

27. Sherri Rose. Mortality Risk Score Prediction in an Elderly Population Using Machine Learning. American Journal of Epidemiology, 177(5):443–452, 01 2013.

28. Romain Pirracchio, Maya L Petersen, Marco Carone, Matthieu Resche Rigon, Sylvie Chevret, and Mark J van der Laan. Mortality prediction in intensive care units with the super icu learner algorithm (sicula): a population-based study. The Lancet Respiratory Medicine, 3(1):42–52, 2015.

29. S. Cohen. Design strategies and innovations in the medical expenditure panel survey. Medical Care, 41: III-5–III-12, 2003.

30. Joel W Cohen, Steven B Cohen, and Jessica S Banthin. The medical expenditure panel survey: a national information resource to support healthcare cost research and inform policy and practice. Medical care, 47(7 Suppl 1): S44—50, July 2009.

31. Jeffrey G Jarvik, Bryan A Comstock, Brian W Bresnahan, Srdjan S Nedeljkovic, David R Nerenz, Zoya Bauer, Andrew L Avins, Kathryn James, Judith A Turner, Patrick Heagerty, Larry Kessler, Janna L Friedly, Sean D Sullivan, and Richard A Deyo. Study protocol: the back pain outcomes using longitudinal data (bold) registry. BMC musculoskeletal disorders, 13:64, May 2012.

32. David Benkeser, Maya Petersen, and Mark J. van der Laan. Improved small-sample estimation of nonlinear cross-validated prediction metrics. Journal of the American Statistical Association, 115(532):1917–1932, 2020.

33. Yu, K., Lu, Z., & Stander, J. (2003). Quantile Regression: Applications and Current Research Areas. Journal of the Royal Statistical Society. Series D (The Statistician), 52(3), 331–350.

34. David Benkeser, Weixin Cai, and Mark J. van der Laan. Rejoinder: A Nonparametric Superefficient Estimator of the Average Treatment Effect. Statistical Science, 35(3):511 –517, 2020.

35. Mark J. Van Der Laan and Sandrine Dudoit. Unified cross-validation methodology for selection among estimators and a general cross-validated adaptive epsilon-net estimator: Finite sample oracle inequalities and examples, 2003.

36. Laan Mark J. van der, Dudoit Sandrine, and Vaart Aad W. van der. The cross-validated adaptive epsilon-net estimator. Statistics & Risk Modeling, 24(3):1–23, December 2006.

37. Partha Deb and Edward C. Norton. Modeling health care expenditures and use. Annual Review of Public Health, 39(1):489–505, 2018. PMID: 29328879.

38. Kathryn P Glass and Jeffery R Anderson. Relative value units: from a to z (part i of iv). The Journal of medical practice management: MPM, 17(5):225—228, 2002.

39. Savannah L. Bergquist, Gabriel A. Brooks, Nancy L. Keating, Mary Beth Landrum, and Sherri Rose. Classifying lung cancer severity with ensemble machine learning in health care claims data. In Finale Doshi-Velez, Jim Fackler, David Kale, Rajesh Ranganath, Byron Wallace, and Jenna Wiens, editors, Proceedings of the 2nd Machine Learning for Healthcare Conference, volume 68 of Proceedings of Machine Learning Research, pages 25–38, Boston, Massachusetts, 18–19 Aug 2017. PMLR.

40. Cheng Ju, Mary Combs, Samuel D. Lendle, Jessica M. Franklin, Richard Wyss, Sebastian Schneeweiss, and Mark J. van der Laan. Propensity score prediction for electronic healthcare databases using super learner and high-dimensional propensity score methods. Journal of Applied Statistics, 46(12):2216–2236, 2019.

